# Synonymous substitution rate slowdown preceding the emergence of SARS-CoV-2 variants and during persistent infections

**DOI:** 10.64898/2026.01.26.26344861

**Authors:** Jennifer L. Havens, Karthik Gangavarapu, Jade C. Wang, Faten Taki, Elizabeth Luoma, Jonathan E. Pekar, Helly Amin, Steve Di Lonardo, Enoma Omoregie, Scott Hughes, Kristian G. Andersen, Tetyana I. Vasylyeva, Marc A. Suchard, Joel O. Wertheim

## Abstract

The emergence of variants has shaped the COVID-19 pandemic. The lack of directly observed precursors to these variants has led to proposals that variants emerge from either persistent infections, transmission in non-human animal populations after reverse-zoonosis, or cryptic transmission in the human population. We investigated the origin of variants by analyzing the molecular clock and rate of nonsynonymous and synonymous substitutions in SARS-CoV-2 circulating in human population, persistently infected individuals, non-human animals, and along variant stems: the branches preceding emergence of SARS-CoV-2 variants (Alpha, Beta, Gamma, Delta, Epsilon, Iota, B.1.637, Mu, and Omicron: BA.1, BA.2/BA.4/BA.5). Along the variant stems we find evidence for an acceleration in the non-synonymous substitution rate, as compared with non-synonymous substitution rate along the branches that represent the genetic diversity of circulating virus. We also find evidence for a slowdown in the synonymous substitution rate preceding the emergence of multiple named variants (e.g., Beta, Delta, Iota, Mu, Omicron BA.1); a similar pattern was observed in some individuals with persistent infections, suggesting that the viral replication rate can slow down during persistent infection. However, the synonymous rate slowdown was not observed for all variants, with some exhibiting an increase in synonymous substitution rates preceding their emergence compared with typical viral transmission (e.g., Alpha, Epsilon). The similarity in evolutionary dynamics preceding some variant emergence and during persistent infections supports the hypothesis that persistent infections were the likely source of many COVID-19 variants.

## Introduction

SARS-CoV-2 emerged in late-2019 and quickly spread across the world. In the ensuing years, lineages with increased propensity for transmission and/or immune escape emerged and were classified as variants of concern and variants of interest. The first named variant, Alpha (B.1.1.7), descended from an early circulating lineage, which we term B.1-like virus. Alpha was characterized by high transmissibility relative to B.1-like virus, largely attributed to N501Y mutation in the spike protein. (Hill et al., 2022; Washington et al., 2021). Starting in late-2020 additional variants with greater transmissibility or immune evasiveness—relative to B.1-like virus—arose, including Beta (B.1.351), Gamma (P.1), Epsilon (B.1.427/B.1.429), Iota (B.1.526), B.1.637, and Mu (B.1.621) (Deng et al., 2021; Faria et al., 2021; Laiton-Donato et al., 2021; Tegally et al., 2021; West et al., 2021). In late 2020, Delta (B.1.617.2) rose to prominence and by mid-2021 became the predominant lineage globally. Shortly thereafter, Omicron (including lineages BA.1, BA.2, BA.4, and BA.5) led to a wave of cases and global weekly infections rose 71% in the last week of December 2021 (Elliott et al., 2021; Lucas et al., 2021; McCrone et al., 2022; Tan et al., 2022; Taylor, 2022; Tegally et al., 2022). The succession of new variants influenced SARS-CoV-2 epidemiology and transmission dynamics, increasing the spread of SARS-CoV-2, as well as the selection of public health response strategies. However, the processes leading to variant emergence were never directly observed. Therefore, understanding the evolutionary dynamics preceding variant emergence is of great public health importance, as it can inform surveillance and prevention efforts.

The Alpha, Beta, Gamma, and Mu variant clades are characterized by a long branch preceding their appearance in the phylogenetic tree, representing substantial divergence relative to previously circulating viruses. Similarly, Epsilon, Delta, Iota, and B.1.637 clades are each characterized by a set of branches, without observed intermediates, that form the stem. The variant “stem branches” tend to have more nonsynonymous substitutions than expected given the rate of evolution associated with B.1-like and within-variant clade virus evolution (Deng et al., 2021; Elliott et al., 2021; Faria et al., 2021; Hill et al., 2022; Laiton-Donato et al., 2021; Lucas et al., 2021; McCrone et al., 2022; Tan et al., 2022; Tegally et al., 2022; Washington et al., 2021; West et al., 2021). Although the positive selection and nonsynonymous substitutions on the stem branches of variants have been repeatedly noted, there has only been limited exploration of the behavior of synonymous and non-coding substitutions (Neher, 2022).

SARS-CoV-2 is generally an acute infection, reaching peak viral load within 4 days, and viral clearance typically within 10 days (Jones et al., 2021; Kissler et al., 2023). However, persistent infections lasting longer than 30 days, sometimes lasting longer than a year, have been documented. In a community surveillance study, an estimated 0.7–3.5% of infections became persistent for longer than 30 days (Ghafari et al., 2024a), often associated with individuals who are immunosuppressed (Corey et al., 2021; Gonzalez-Reiche et al., 2022; Machkovech et al., 2024; Scherer et al., 2022). Some persistent infections are characterized by accelerated evolution resulting in many nonsynonymous mutations that allowed the virus to evade the host immune response (Ghafari et al., 2024b; Machkovech et al., 2024). The stem branches of variants have the same pattern of accelerated nonsynonymous substitutions, and so persistent infections have been proposed as a possible source of variants. It has also been proposed that variants emerge from transmission in non-human animal populations after reverse-zoonosis and cryptic transmission in the human population (Kupferschmidt, n.d.; Markov et al., 2023; Shrestha et al., 2022; Sparrer et al., 2023; Tegally et al., 2022; Wei et al., 2021). Evolution in non-human animals has been suggested because the virus can undergo rapid evolution, which could be unobserved in these undersampled non-human animals (Sparrer et al., 2023; Wei et al., 2021). Cryptic transmission in human populations with poor genomic surveillance could result in long branches as evolution occurs unobserved due to biased undersampling (Shrestha et al., 2022; Tegally et al., 2022).

In this study, we compare the evolutionary dynamics in variant stem branches, persistent infections, and non-human animal hosts with evolution during typical transmission between humans. We consider the stem branches of the Alpha, Beta, Gamma, Delta, Epsilon, Iota, B.1.637, and Omicron variants. Using a Bayesian phylodynamic framework with local molecular clocks, we demonstrate how changes in the rate of synonymous substitutions point to the mechanism behind variant emergence.

## Results

To contextualize the origin of SARS-CoV-2 variants, we assembled genomic datasets representing four modes of SARS-CoV-2 evolution: typical human-to-human transmission, long-term persistence within individuals, transmission between non-human animals following reverse zoonosis, and the phylogenetic stem branches preceding emergence of variants.

### SARS-CoV-2 variant stem branches

We identified the stem branches preceding the B.1 variants (Beta, Delta, Epsilon, Mu, Iota, and B.1.637) and B.1.1 variants (Alpha, Gamma, Omicron) on a maximum likelihood phylogenetic tree with 10,572 high quality SARS-CoV-2 genomes representing circulating genomic diversity in New York City (NYC), over a 2 year period (**Fig 1**). We focused on NYC because it is a major urban center with robust public health surveillance that produced a large volume of high-quality surveillance of SARS-CoV-2 genomes reflecting global viral diversity, including all variants of interest. Branches that were not associated with a variant stem were considered to represent evolution associated with typical human-to-human transmission, including both within variant clades and all branches that preceded the evolution of variants (such as B.1 and B.1.1 virus), which we term B.1-like virus. For each variant, we compared the evolution on the stem branches to evolution within B.1-like phylogeny and the variant clade. For each variant (**Fig 1**), we applied local molecular clocks on variant stem branches in a Bayesian phylodynamic inference framework.

**Fig 1.**
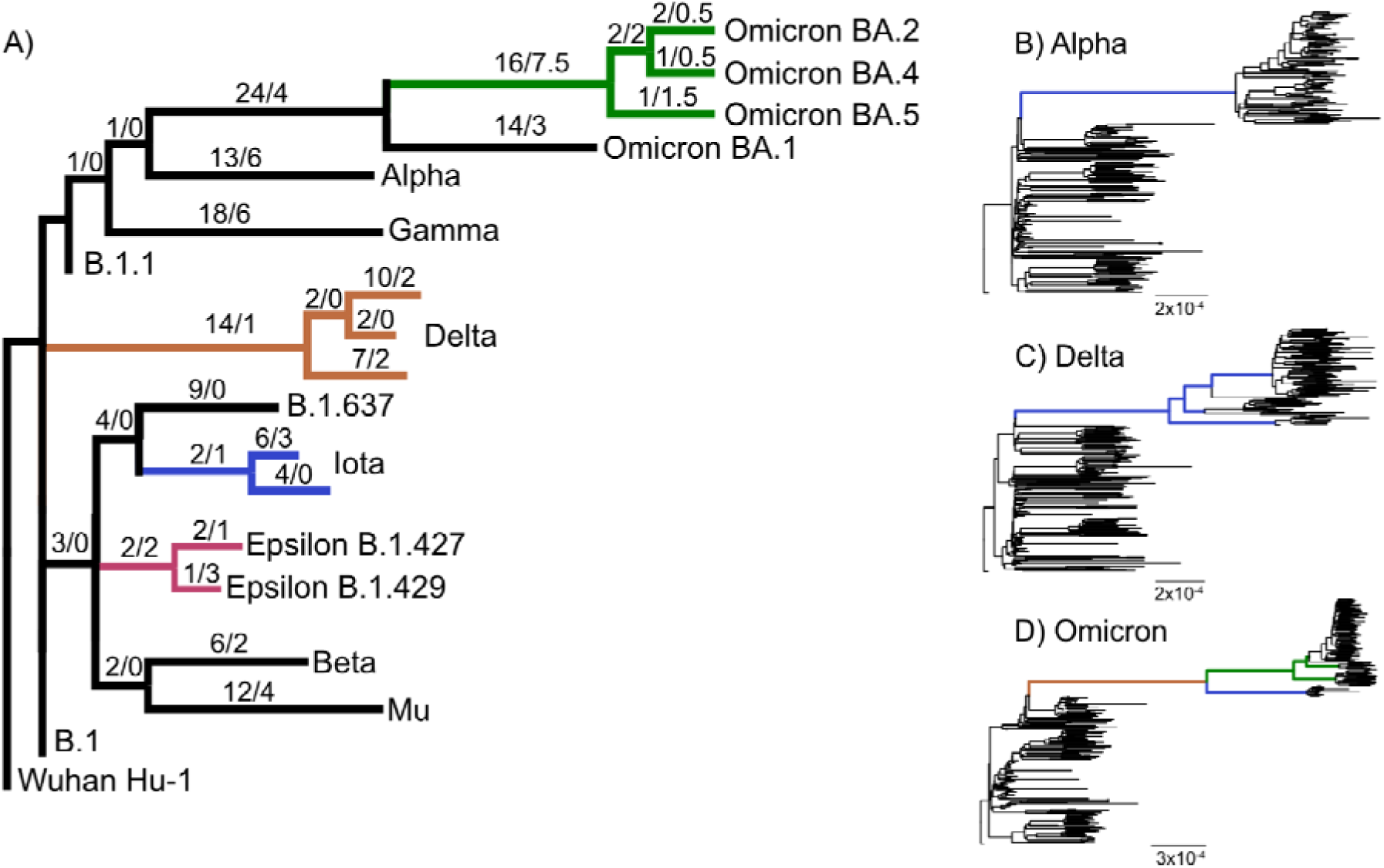
Phylogenetic trees of variant stem branches. A) Cartoon tree showing SARS-CoV-2 backbone and the relationship between variants, annotated with substitutions formatted as the number of nonsynonymous/synonymous substitutions inferred by robust counting in BEAST. B) Phylogenetic tree of B.1-like and Alpha sequences, the variant stem branch is highlighted in blue. C) Phylogenetic tree of B.1-like and Delta sequences, the variant stem branches is highlighted in blue. D) Phylogenetic tree of B.1-like and Omicron sequences, BA.1 stem branch is highlighted in blue, the stem branches leading to BA.2, BA.4, and BA.5 (BA.2/BA.4/BA.5) is highlighted in green, the base of Omicron, which included in analysis the branches for BA.1 and BA.2/BA.4/BA.5, is highlighted in brown.

### Identification of persistent SARS-CoV-2 infections

To identify persistent SARS-CoV-2 infections, we inferred a phylogenetic tree from viral genomes sampled from individuals with two or more genomes sampled >30 days apart in New York City (genomes n=396, individuals n=194). We defined persistent infections as instances in which viral genomes sampled from the same individual were monophyletic. This approach yielded persistent infections that were contemporaneous to the stem branches of the named variants with consistent location and sequencing approach between all the genomes. We found 15 individuals with persistent infections, represented by 46 viral genomes. The remaining 181 individuals were determined to most likely be instances of re-infection. Seven people with persistent infections had viral evolution comprising more than 3 person months of evolution across the phylogeny; thus we only included these seven individuals in the subsequent analyses. These viral genomes were B.1-like (n=2), Alpha (n=1), Delta (n=2), Iota (n=1), and B.1.637 (n=1). We applied a local molecular clock for viruses from each individual with persistent infection in Bayesian phylodynamic inference (**Fig 2**) and compared the evolution during these persistent infections with evolution within their respective variants.

**Fig 2.**
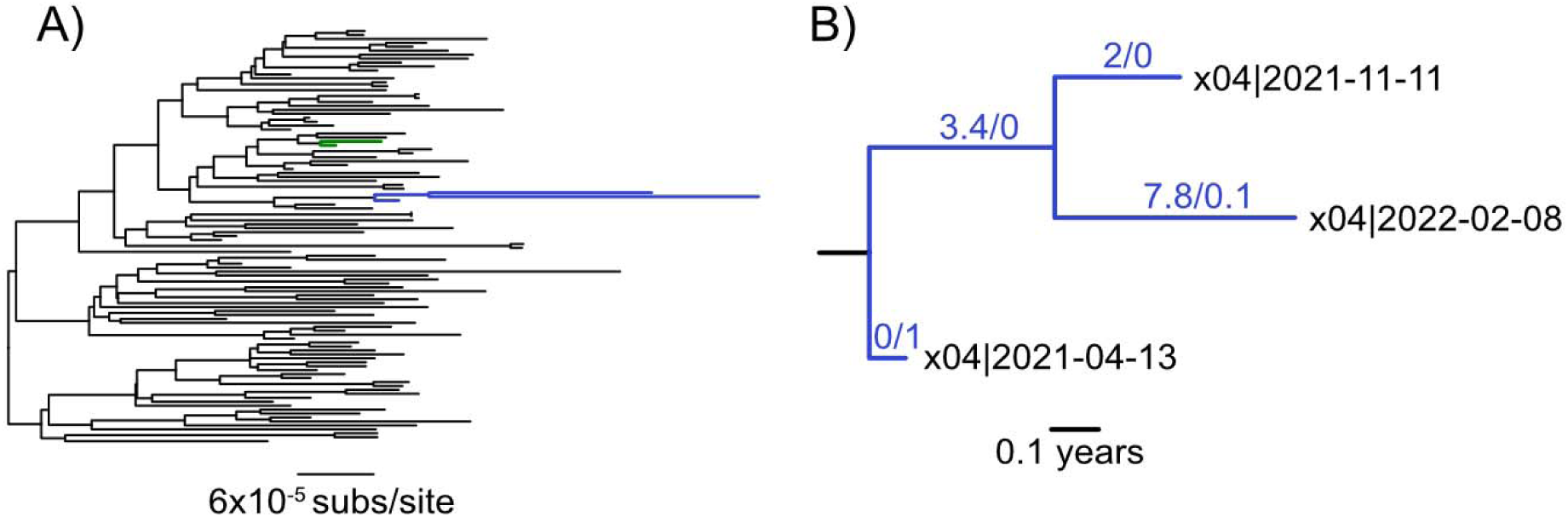
Phylogenetic trees of persistent infections. A) Phylogenetic tree of Alpha sequences, persistent infection clades are highlighted in blue (Alpha_x04) and green. B) Phylogenetic tree of persistent infection Alpha_x04 with time scaled branches annotated with robust counts of nonsynonymous/synonymous substitutions.

### SARS-CoV-2 clades in deer

To characterize SARS-CoV-2 evolution in non-human animals after reverse zoonosis, we used 22 viral genomes from clades of deer SARS-CoV-2 arising from 5 previously characterized reverse zoonosis events (McBride et al., 2023). All deer virus samples were descended from Alpha and Delta variants. We applied a local molecular clock for each deer clade in Bayesian phylodynamic inference in the phylogenetic context related to human virus sequences from their respective Alpha and Delta variants.

### SARS-CoV-2 synonymous substitution rate

We inferred the rate of synonymous substitutions over time (i.e., synonymous substitutions across all coding regions/year) using a robust counting method (Lemey et al., 2012; O’Brien et al., 2009) in a phylodynamic framework (Suchard et al., 2018). Along each partition (i.e., variant stem branches, persistent infection clades, deer clades, and background virus), the rate of substitutions was calculated as the sum of inferred synonymous substitutions divided by the cumulative branch length. The inferred background rate of synonymous substitutions along internal branches in coding regions of B.1-like virus was 6.8 substitutions/year [95% highest posterior density (HPD): 6.1–8.2 substitutions/year] (**Supp Table 1**).

**Table 1.**
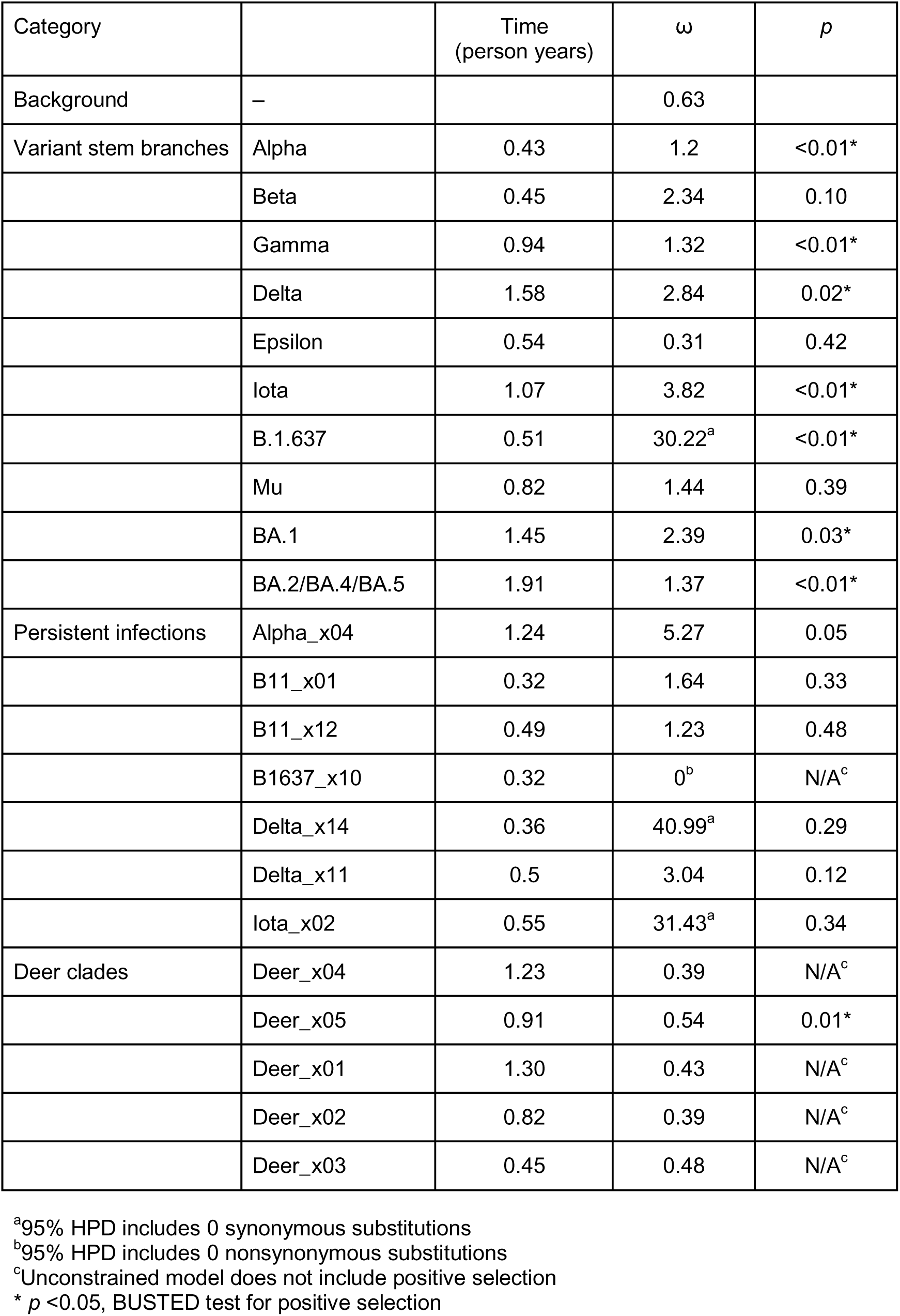
Estimate of ω and evidence for positive directional selection.

### Synonymous substitution rate preceding variants

We then compared the synonymous substitutions rate on the stem branches preceding variants with the synonymous substitutions rate during typical B.1 transmission and within the corresponding variant clade. We observed a pattern along the stem branches leading to Beta, Delta, Iota, B.1.637, Mu, and BA.1, whereby the rate of synonymous substitutions was significantly slower than the inferred background synonymous rate (**Fig 3A**). For example, the stem branches of the Delta variant encompass 1.6 person years, with an expectation of approximately 11 synonymous substitutions (95% HPD: 9.5–12.8); however, we only observed 5 synonymous substitutions (**Supp Table 2**), less than half of the amount expected. The synonymous substitution rate was 3.2 substitutions/year (95% HPD: 2.8–3.8 substitutions/year). Similarly for the Iota variant, we observed only 4 synonymous substitutions (**Supp Table 2**) over the 1.1 person-years comprising the variant stem branches; in contrast, under the synonymous substitutions rate during human-to-human transmission, we would have expected 7 synonymous substitutions (95% HPD: 6.0–9.2 substitutions) over this time period. We observed a synonymous substitution rate of 3.8 substitutions/year (95% HPD: 2.9–5.1 substitutions/year) along the Iota stem branches. Similarly in the stem of BA.1 the expected number of synonymous substitutions is 10.2 (95% HPD: 9.2–11.2 substitutions), however only 7 synonymous substitutions are observed on the BA.1 stem branches.

**Fig 3.**
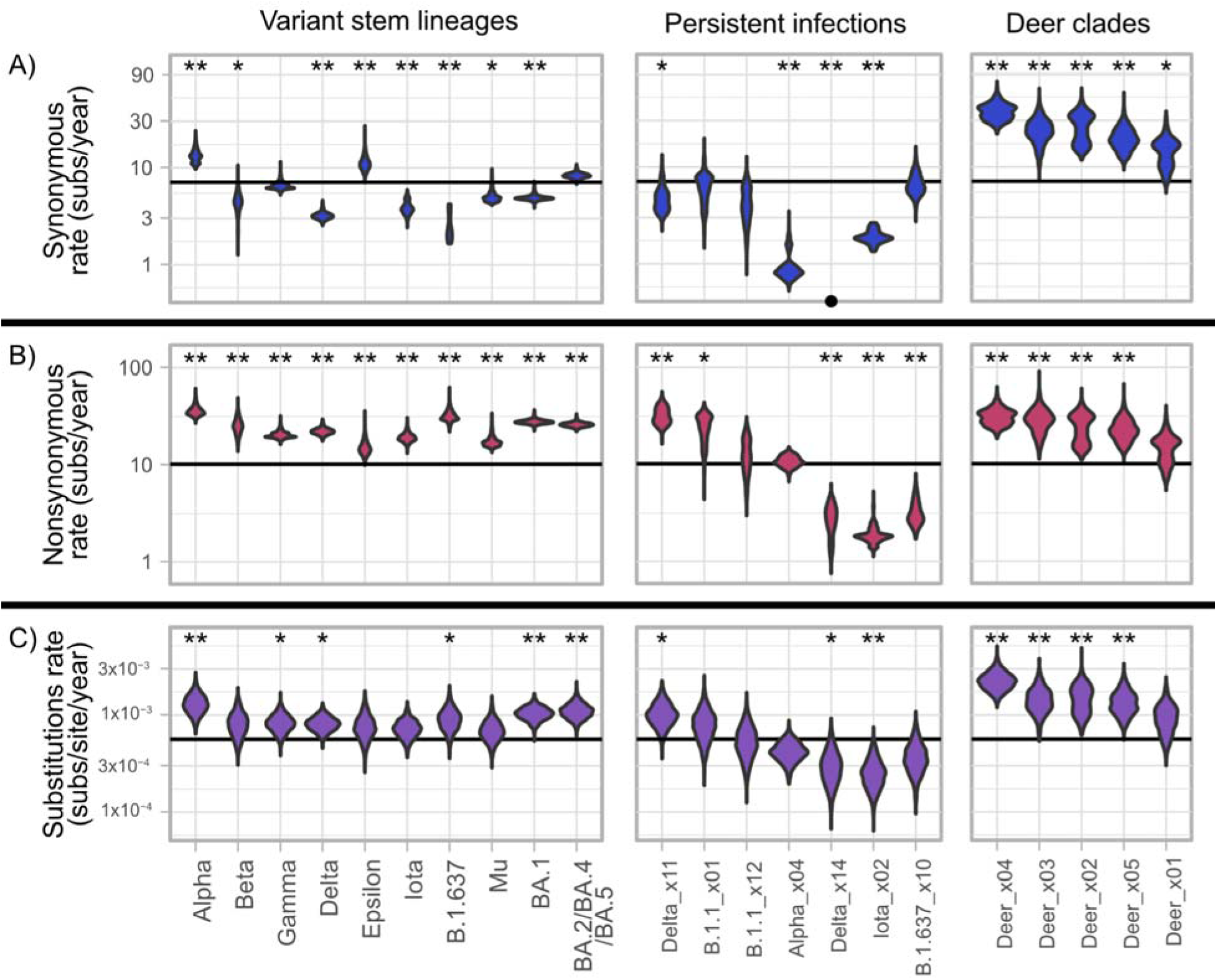
Rate of nonsynonymous and synonymous substitutions and the molecular clock. A) Summary of molecular clock rate for each variant stem branches, persistent infection, and deer clade with posterior probability that the rate is larger or smaller than the background rate (* P >0.9; ** P >0.99). Background molecular clock median (5.6x10^-4^ substitutions/site/year) indicated by black horizontal line. B) Summary of nonsynonymous substitution rate with posterior probability that the rate is larger or smaller than the background rate (* P >0.9; ** P >0.99). Background nonsynonymous rate median (10.1 substitutions/year) indicated by black horizontal line. C) Summary of synonymous substitution rate with posterior probability that the rate is larger or smaller than the background rate (* P >0.9; ** P >0.99). Background synonymous rate median (6.8 substitutions/year) indicated by black horizontal line.

In contrast to most variants we examined, there is evidence of increased rate of synonymous substitutions in the Alpha and Epsilon stem branches (**Fig 3A**). For example, the Alpha stem branch represents 0.43 person years of evolution and has 6 synonymous substitutions, double the expected 3.0 synonymous substitutions (95% HPD: 2.3–3.5 substitutions). The Alpha stem branch had a rate of 13.1 synonymous substitutions/year (95% HPD: 10.0–17.7 substitutions/year).

We did not detect an increase or decrease in the synonymous substitution rate on the Gamma or BA.2/BA.4/BA.5 stem branches (**Fig 3A**). We inferred a rate of 6.4 substitutions/year (95% HPD: 5.6-7.9 substitutions/year) leading to Gamma and 8.3 substitutions/year (95% HPD: 7.3–9.2 substitutions/year) leading to BA.2/BA.4/BA.5, which is consistent with the background rate of 6.8 synonymous substitutions/year (95% HPD: 6.1–8.2 substitutions/year).

### Synonymous substitution rate during persistent infections

When we considered persistent infections, 4 of the 7 persistent infections had a slower rate of synonymous substitutions compared with the background (**Supp Table 1 & Fig 3A**). For example in the case of Alpha_x04, there was 1 synonymous substitution over 1.2 person years and the synonymous rate was 0.8 substitutions/year (95% HPD 0.6–1.8 substitutions/year). This rate was substantially slower than the synonymous substitution rate of 6.8 synonymous substitutions/year in the context of human-to-human transmission. This slowdown in synonymous substitution rate is comparable to that seen in the Beta, Delta, Iota, B.1.637, Mu, and BA.1 variant stem branches.

In 3 of the 7 persistent infections, the rate of synonymous substitutions was not significantly different from the background (**Fig 3A**). For example, B.1.637_x10 had 2 synonymous substitutions over 0.32 person years and a synonymous substitution rate of 6.4 substitutions/year (95% HPD: 3.9–10.5 substitutions/year), which is consistent with the synonymous substitutions rate expected during typical transmission.

### Synonymous substitution rate in deer virus

We also compared typical transmission to evolution within clades of SARS-CoV-2 sampled in deer. We found that there was an increased rate of synonymous substitutions in all 4 deer clades compared with human-to-human transmission (**Fig 3A**). For example, deer clade Deer_x05 had 17 synonymous substitutions over 0.91 deer infection years and a rate of 19.1 synonymous substitutions/year (95%HPD: 11.4–30.2), which is faster than expected during transmission within human hosts.

### Nonsynonymous substitution rate

We inferred the rate of nonsynonymous substitutions over time (i.e., nonsynonymous substitutions across all coding regions/year) using the same approach used to infer the rate of synonymous substitutions. Using B.1-like virus genomes to represent typical human-to-human transmission we inferred a background rate of 10.1 nonsynonymous substitutions/year (95%HPD: 9.1–11.2 substitutions/year).

All variant stem branches that we considered had an elevated rate of nonsynonymous substitutions relative to the background (**Fig 3B; Supp Table 1**). For example, the Alpha stem branch had a rate of 35.5 nonsynonymous substitutions/year (95% HPD: 29.7–45.6 substitutions/year) with 15 nonsynonymous substitutions over 0.43 person years (**Fig 1**). The stem branches of Iota which had 22 nonsynonymous substitutions over 1.1 person years with a rate of 19.0 nonsynonymous substitutions/year (95% HPD: 15.3–23.6 nonsynonymous substitutions/year), which exceeds the expected rate of nonsynonymous substitutions. The excess number of nonsynonymous substitutions that has been previously described (Hill et al., 2022; West et al., 2021).

In persistent infections, 2 of 7 had an increased rate of nonsynonymous substitutions compared with the rate associated with human-to-human transmission. One of these persistent infections of increased rate was Delta_x11, which had a rate of 31.2 nonsynonymous substitutions/year (95% HPD: 22.3–46.2 substitutions/year). This increased rate is consistent with the dynamics observed on variant stem branches. We observed high variance between nonsynonymous substitution rates in persistent infections, consistent with previous findings (Ghafari et al., 2024b). Of 7 persistent infections, 3 had a slower rate of nonsynonymous substitutions compared with rates under human-to-human transmission. For example, Iota_x02 had 1 nonsynonymous substitution in 0.55 person years and a nonsynonymous substitution rate of 1.8 nonsynonymous substitutions/year (95% HPD: 1.3–2.6 substitutions/year). The remaining 2 persistent infections we considered had a similar rate of nonsynonymous substitutions compared to the background (n=2). For example, Alpha_x04 had a rate of 10.7 nonsynonymous substitutions/year (95% HPD: 8.4–13.6 substitutions/year), which is consistent with the rate of nonsynonymous substitutions during typical transmission: 10.1 nonsynonymous substitutions/year.

McBride et al.(McBride et al., 2023) found an increased rate of nonsynonymous substitutions in deer; our approach recapitulated this finding. (**Fig 3B**). For example, the clade Deer_x05 had a rate of 22.4 nonsynonymous substitutions/year (95% HPD: 13.1–35.3 substitutions/year), with 20 nonsynonymous substitutions over 0.91 deer infection years.

### Molecular clock and selection in human-to-human transmission and preceding variants

We considered the interaction of nonsynonymous and synonymous substitutions by estimating the overall molecular clock, which is the nonsynonymous and synonymous rates combined, and the ratio of these rates when normalized [nonsynonymous substitutions per nonsynonymous site (dN) and synonymous substitutions per synonymous site (dS)],dN/dS or ω. The ω measure is a useful and informative statistic describing the nature of selective forces. We inferred ω and tested for positive selection using BUSTED (Murrell et al., 2015), and inferred overall clock rates from the Bayesian analysis. We inferred the background rate of evolution and ω associated with transmission using B.1-like sequences. The overall rate of substitutions in coding regions was 5.5x10^-4^ substitutions/site/year (95% HPD: 5.1x10^-4^ – 6.2x10^-4^ ; **Supp Table 1**), and ω was 0.63.

Variant stem branches tended to have a molecular clock rate that was elevated or comparable to the background clock rate, and stem branch ω was typically greater than the background ω (**Table 1**; **Fig 3C**). The Alpha, Gamma, and the BA.2/BA.4/BA.5 stem branches had an increase in the overall rate and ω, largely driven by higher nonsynonymous substitutions rates (**Fig 3; Supp Table 1**). The stem branches of Beta, Delta, Iota, B.1.637, Mu, and BA.1 variants also had an increase in nonsynonymous rates; however, due to a decrease in the synonymous substitution rates, the overall molecular clocks did not increase as much, compared with the other variants (**Fig 3; Supp Table 1**). The decrease in synonymous rate explains the noted lack of speed up in the overall molecular clock of the Delta stem branches despite the same excess of nonsynonymous substitutions that is seen in the Alpha stem branch (Hill et al., 2022). The increase in ω was driven by both the increase in nonsynonymous and decrease in synonymous substitutions rates (**Table 1**). Although, we note that 95% HPD of the B.1.637 stem branch included zero synonymous substitutions, which makes inference of ω difficult (**Supp Table 2**).

The Epsilon stem branches did not show the same behavior as other variants, as it was the only variant stem branches where ω was not higher than the background. The overall molecular clock rate of the Epsilon stem branches was not significantly different from the background rate, unlike other variant stem branch dynamics.

### Molecular clock and selection during persistent infections

In the 7 persistent infections we analyzed, most had a change in the overall clock rate and ω compared with the background (**Table 1**; **Fig 3C**). Some persistent infections had changes in rates that are similar to that in variant stem branches. For example, B.1.1_x01 had an increased ω, and nonsynonymous substitution rate and a synonymous and overall substitution rate that is consistent with background transmission. This change in rates was largely the same in the Beta, Iota, and Mu stem branches, although these variant stem branches do have a reduced rate in synonymous substitutions. Some persistent infections did have a reduced rate of synonymous substitutions and increase in ω, which is characteristic of many variant stem branches, such as Alpha_x04, Iota_x02 and Delta_x14 (**Fig 3A; Supp Table 2**). Overall we found persistent infections can have the same changes in the molecular clock and ω compared with the background that was characteristic of variant stem branches.

### Molecular clock and selection in deer virus

In the 4 clades of SARS-CoV-2 sampled from deer, we saw a consistent pattern of an increase in nonsynonymous and synonymous substitution rate, and a corresponding increase in the overall rate of substitutions (**Table 1**; **Fig 3**). This increased rate is consistent with previous findings (McBride et al., 2023). The selective pressure across all deer clades, assessed by ω, tended to be similar to the background, because the nonsynonymous and synonymous rates increased relatively proportionally. This finding is consistent with a lack of gene wise positive selection previously described (McBride et al., 2023).

## Discussion

We performed phylodynamic analysis to understand the evolutionary dynamics of the phylogenetic branches ancestral to SARS-CoV-2 variants. We found evidence for a slowdown in the absolute rate of synonymous substitutions preceding the emergence of several variants (Beta, Delta, Iota, and Omicron), accompanied by an increase in the absolute rate of non-synonymous substitutions. We detected this same pattern of evolutionary dynamics in persistent SARS-CoV-2 infections. In contrast, no such slowdown in the synonymous substitution rate was observed during typical transmission between humans or between deer after reverse-zoonosis. These findings strongly support the conclusion that SARS-CoV-2 variants arise during persistent infection and in neither an animal reservoir nor during cryptic human-to-human transmission.

We found some persistent infections have slower rates of synonymous substitutions, as was seen in a subset of variant stem branches. A change in synonymous substitution rate is often associated with a change in viral replication rate (Hanada et al., 2004; Holmes, 2003; Jenkins et al., 2002). The slower rate of evolution could be due to lingering non-replicating and non-viable RNA (Ghafari et al., 2024a); however, we note during continued evolution there are fewer synonymous substitutions than expected on branches across the full Alpha_x04 clade (**Fig 1B**) suggesting continued replication of virus with slower synonymous substitutions. Synonymous substitution rate can be impacted by selection, rate of viral replication, and fidelity in sequence replication (Pond and Muse, 2005).

We observed changes in the nonsynonymous substitutions rate in persistent infection and variant stem branches. The increased rate of nonsynonymous substitutions in variant stem branches has been attributed to selection pressure (Hill et al., 2022; McCrone et al., 2022; Volz et al., 2021). In persistent infections there is greater variance in the nonsynonymous substitution rate than in the synonymous substitutions rate (Ghafari et al., 2024b; Harari et al., 2024). This larger variance in nonsynonymous substitutions can be explained by the disproportionate impact of selection on nonsynonymous substitutions and higher rate of substitutions. Mutations associated with VOCs have been observed arising in persistent infections (Joseph et al., 2025). The high variability in nonsynonymous substitution rate in persistent infections is consistent with variant stem branches, and unlike human-to-human transmission.

We characterized human-to-human transmission dynamics using surveillance sampling because most evolution is associated with evolution during transmission between acute infections. However, we have not censored from our dataset any evolution that could be associated with onward transmission from a persistent infection, or other distinct mechanisms, as long as it was not considered a variant stem branch. The typical human-to-human transmission we characterize likely includes some evolution from persistent infections that were unidentified, as well as human-to-human transmission of acute infections, so our comparisons between human-to-human transmission and persistent infections are conservative in nature due to the diluted inference.

Together the molecular clock, selection dynamics, and deer characteristic mutations suggest that reverse zoonosis to deer is not the primary source of variants. SARS-CoV-2 in deer following reverse zoonosis exhibited an increased rate of both nonsynonymous and synonymous substitutions, and selection pressure similar to human transmission; these selection dynamics are distinct from the dynamics observed on most variant stem branches. Additionally, SARS-CoV-2, circulating in deer, tends to acquire characteristic substitutions which are not seen in variant stem branches (Hale et al., 2022; McBride et al., 2023). We cannot exclude the possibility that infection in another non-human animal could be the source of SARS-CoV-2 variants as we only analyzed SARS-CoV-2 in deer and not other non-human hosts; however, SARS-CoV-2 evolution in deer does not recapitulate the evolutionary dynamics preceding the emergence of variants.

Variant stem branches have different rates of evolution and selection dynamics compared to typical human-to-human transmission, suggesting cryptic transmission in the human population is not the source of SARS-CoV-2 variants. The overall molecular clock of some variant stem branches (e.g., Delta) is consistent with human-to-human transmission, leading to the suggestion that variants like Delta could have emerged from typical transmission (Hill et al., 2022). However, our analysis uncovered that this molecular clock is not consistent with human-to-human transmission when we consider that a slower rate of synonymous substitutions is obfuscating an increase in nonsynonymous substitutions. The rate of synonymous substitutions associated with typical transmission within variant clades has broadly remained unchanged throughout the pandemic (Neher, 2022).

SARS-CoV-2 primarily infects cells in the upper respiratory tract, but can move to other areas of the body, where it experiences distinct selection patterns (El Moussaoui et al., 2024; Jary et al., 2020; Rueca et al., 2020; Van Cleemput et al., 2021). The slowdown in the synonymous substitution rate preceding variants could be a consequence of persistent infection in non-respiratory cells. Synonymous substitution rate and viral replication can be affected by host species (Kim et al., 2022), cell tropism (Hicks and Duffy, 2014) and virus sequestration (Keita et al., 2021). In HIV, the synonymous substitution rate varies between infections and impacts the dynamics of the infection (Lemey et al., 2007). Understanding how SARS-CoV-2 evolves in different areas of the body will require more samples from the diverse body compartments. Viral genomics from wastewater suggests that SARS-CoV-2 experiences distinct patterns of evolution in gut tissue; however, the vast majority of SARS-CoV-2 genomes available for analysis are from the upper respiratory tract (Machkovech et al., 2024).

A clue to the slowdown in synonymous substitution rates may be found in another virus. Ebola virus, although typically causing an acute infection, can persist in immunoprivileged sites where it underwent substantially slower rates of replication (Whitmer et al., 2018). Transmission after long-term persistence has seeded multiple Ebola outbreaks, including those where the stem branch leading to the outbreak had fewer synonymous substitutions than expected given the amount of time of evolution (Keita et al., 2021).

SARS-CoV-2 can replicate in different body compartments (Van Cleemput et al., 2021) and, like Ebola virus, produce persistent infections exhibiting slower rates of synonymous substitutions. These observations, combined with the dearth of synonymous substitutions preceding the emergence of SARS-CoV-2 variants, suggests that SARS-CoV-2 may experience varied replication rates in additional anatomical compartments during persistent infection, which may ultimately be the source of SARS-CoV-2 variants. The likely contribution of persistent infections in variant emergence highlights the importance of further research into the dynamics and prevention of persistent SARS-CoV-2 infections.

## Methods

### Data availability

Consensus genomes and analysis used is available in XMLs (**Supp Data 1-18**). Consensus genomes or sequencing data is available for query for all samples through NCBI, GISAID, and SRA. IDs of samples used for B.1-like and variant characterization are available in **Supp Table 3**. IDs and matching information for persistent infection sequences are available in **Supp Table 4**.

### SARS-CoV-2 genome surveillance

We analyzed SARS-CoV-2 full genome sequences (<1% Ns) from New York City Public Health Laboratory (PHL) collected from 7 February 2020 through 1 June 2022 (n=10,572), previously deposited in GenBank and SRA. We used data from PHL alone because the consensus genome sequences are high quality, the metadata has high fidelity, information on repeated tests is available, and all variant stem branches can be sufficiently characterized even using this subset of global data. Pangolin lineages (v4.1.2) were assigned using UShER (O’Toole et al., 2021). Genomes were aligned with MAFFT (v7.486) (Katoh et al., 2005) to the reference genome Wuhan Hu-1, and coding regions were extracted for further analyses.

We used a random subset of 150 genomes from all genomes not assigned to a Pangolin variant to represent the early background context (B.1-like) (accession numbers are reported in **Supp table 3**). We selected smaller datasets to accommodate the Bayesian Robust Count models (see below). Using BEAST (v1.10.5) (Suchard et al., 2018) we inferred a molecular clock rate for these early sequences using only the coding regions of 5.4x10^-4^ substitutions/site/year (95% HPD: 4.9x10^-4^ – 5.9x10^-4^ substitutions/site/year).

### Characterizing SARS-CoV-2 variant stem branches

For each variant, we used the B.1-like background sequences and all variant sequences to infer a tree with IQTREE (v1.6.12) (accession numbers reported in **Supp table 3**) (Minh et al., 2020). Using these trees we defined a variant stem as the path between B.1 or B.1.1 to the base of the variant polytomy expansion (**Fig 2A**). For Alpha, Beta, B.1.637, Gamma, and Mu the variant stem was a single branch (**Fig 1B, Supp Fig 1-5**). The Epsilon variant stem is the branch leading to B.1.427, the branch leading to B.1.429 and their parent branch (**Supp Fig 6**). Delta and Iota variant stem branches are the set of branches preceding the polytomy expansion that is characteristic of variants (**Fig 2C,D, Supp Fig 7,8**). We subset the Omicron stem branches into two sets for defining the variant stems (BA.1 alone and BA.2, BA.4, and BA.5 combined), but included all Omicron sequences in one alignment for inference. The stem branch of BA.1 is the branch leading to the most recent common ancestor (MRCA) of BA.1, BA.2, BA.4, and BA.5 and the branch leading to the MRCA of BA.1 (**Fig 2D, Supp Fig 9**). The stem of BA.2, BA.4 and BA.5 is multiple branches leading from the MRCA with BA.1 to the MRCA of each lineage (BA.2, BA.4, BA.5). We chose to combine BA.2, BA.4, and BA.5 because they were closely related and some sites showed repeated changes without observed intermediates (S:452). The base stem for Omicron was included in both the BA.1 stem and the BA.2, BA.4, and BA.5 stem because it represented significant divergence that we felt it was important to include, but could not be reasonably attributed to only BA.1 or BA.2, BA.4, and BA.5 alone.

We note that because each variant stem is defined without other variant sequences there is some shared evolutionary history, i.e. from the base of the B.1 polytomy to Beta, Epsilon, Iota, Mu, and B.1.637, and from the bases of B.1.1 to Alpha, Gamma, and Omicron. However, based on ancestral reconstruction less than 20% of substitutions of any variant are shared with another variant stem (**Figure 1A**), with exception of the Omicron variants sharing a base (Figure 1D) and B.1.637 and Iota sharing more. Delta does not have shared evolutionary history with any other stem.

### Persistent intra-host infection phylodynamics

To identify persistent infections, we used all genomes from individuals with more than 2 SARS-CoV-2 genomes sampled more than 30 days apart by New York City Public Health Laboratory, NYC Pandemic Response Laboratory, or Department of Health Wadsworth Center (n=373). We inferred a phylogenetic tree using IQTREE using the GTR+F+G_4_ substitution model, retaining polytomies, and enforcing a minimum branch length of 10^-9^ substitutions/site, and identified persistent infections as individuals that had multiple genomes that formed a cluster (samples could be connected without passing through a node representing a sample from another individual). By using genomes from NYC surveillance, we ensured that persistent infections that are contemporaneous to the variant stems of interest had consistent location and sequencing approach between them; additionally this approach mitigates overrepresentation from immune compromised individuals often seen in convenience sampling. We found a total of 15 individuals with persistent infections, characterized by 42 sequences (accession numbers are reported in **Supp Table 4**). For each case we inferred a local clock with BEAST, using sequences of the same variant, or the B.1-like subset, as context (**Supp Fig 10-14**).

### Deer virus genomes

To investigate SARS-CoV-2 evolution associated with a deer host we considered 5 clades, comprising 22 previously published genomes (McBride et al., 2023). We choose clades with more than 0.33 years of evolution inferred in previous analysis (**Supp Fig 15, 16**).

### Bayesian local clock inference and robust counting

We inferred a local molecular clock for each variant stem, persistent infection, and deer clade using BEAST with a GTR for each codon position (**Supp Data 1-18**). For each variant stem we used the B.1-like background sequences for context together with all variant sequences. For persistent infections and deer clades, all sequences of the same variant from PHL were used for context. Context sequences (variant and B.1-like) were given a tight molecular clock prior (5.4x10^-4^ substitutions/site/year, standard deviation: 2.3x10^-5^), inferred from the B.1-like context. We inferred a local clock on the stem of the variant clades, and within the persistent infection and deer clades. For Omicron, the base branch (parent branch of BA.1 and BA.2 BA.4 and BA.5) shared a local clock with the BA.1 stem, while the stem of BA.2 BA.4 and BA.5 was an independent local clock. We included the base branch for both stems in the post-BEAST analysis. The local clock was set as the product of the background rate and an increment factor with a log normal prior centered on 1. For each analysis we ran 2 chains for 5x10^8^ to 10^9^ states and investigated the chain convergence using Tracer (v1.7.2) (Rambaut et al., 2018), then used LogCombiner (v10.5.0) (Suchard et al., 2018) to resample and combine the obtained tree distributions at a lower frequency. We used robust counting (O’Brien et al., 2009) to infer the amount of synonymous and nonsynonymous substitutions on branches. We did not consider insertions and deletions or noncoding regions.

For each set of branches of interest we summarized the person years (i.e., sum of the length of branches of interest), clock rate, synonymous substitution rate, and nonsynonymous substitution rate as compared to the background context. The posterior probability that the rate is larger or smaller than the background rate is calculated by counting the number of states in which the local rate is greater than the background for each tree in the posterior, and taking the maximum of the proportion of states that are greater, or one minus that proportion (e.g., the minimum posterior is 0.5). In our interpretation, we considered only variant stems, persistent infections, and deer clades with more than 0.25 person years. The variant under monitoring B.1.1.519 and 5 persistent infections were excluded due to this criteria.

### Inference of **ω** and positive selection

Using BUSTED (v2.5.42) (Murrell et al., 2015), we inferred the ω or dN/dS, and tested for positive selection on the branches of interest described above on the same phylogenetic trees. All other branches were considered part of a nuisance set.

## Data Availability

Consensus genomes or sequencing data is available for query for all samples through NCBI, GISAID, and SRA. IDs of samples used for B.1-like and variant characterization are available in Supp Table 3. IDs and matching information for persistent infection sequences are available in Supp Table 4.

## Supplementary information

**Supp Table 1. Molecular clock and rate of nonsynonymous and synonymous substitutions**

**Supp Table 2: Synonymous substitutions of variant stem branches**

**Supp Table 3. Ascension numbers and ID of context sequences**

**Supp Table 4. Ascension numbers and ID of persistent infection sequences**

## Acknowledgements

This work was partially supported through US National Institutes of Health grants R01 AI135992 (J.L.H, J.O.W.), U19 AI135995 (K.G., K.G.A., M.A.S.), and R01 AI153044 (J.L.H., M.A.S.). J.E.P. acknowledges support from NIH (T15LM011271), the UC San Diego Merkin Fellowship, and The Rockefeller Foundation (PC-2022-POP-005).

