## Supplementary figures and images for "Synonymous substitution rate slowdown preceding the emergence of SARS-CoV-2 variants and during persistent infections"

### Supp Fig 1

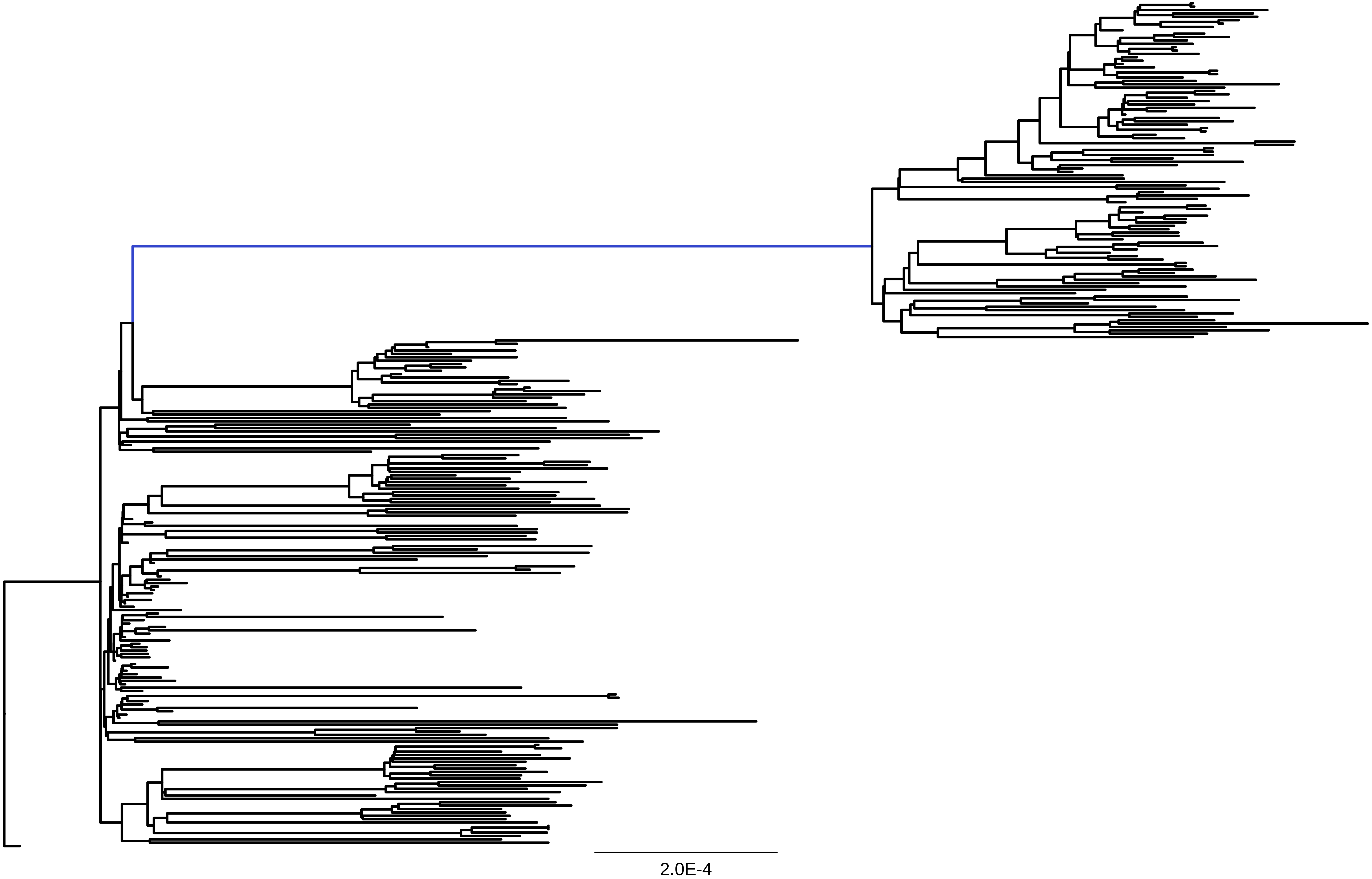

### Supp Fig 2

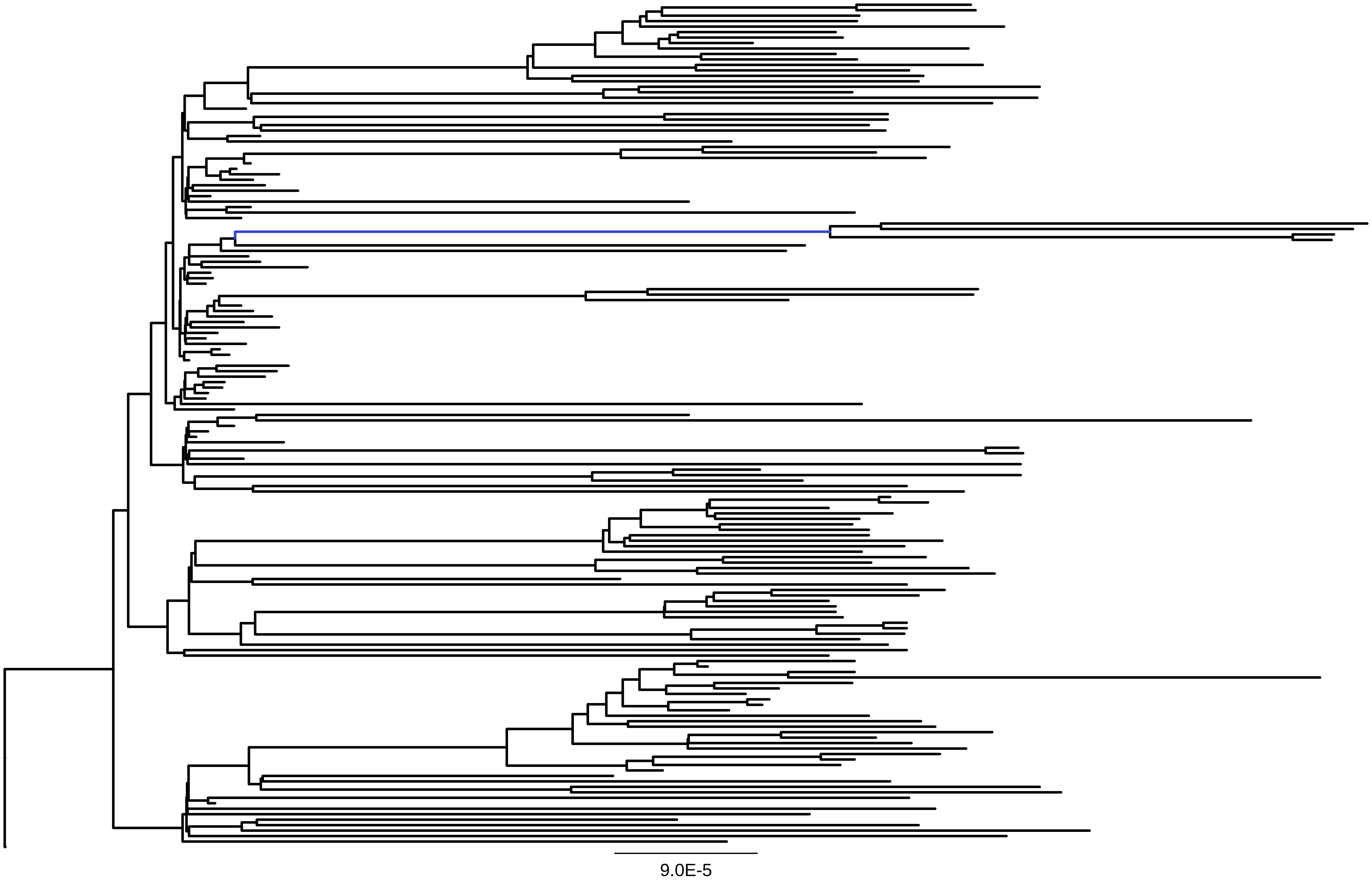

### Supp Fig 3

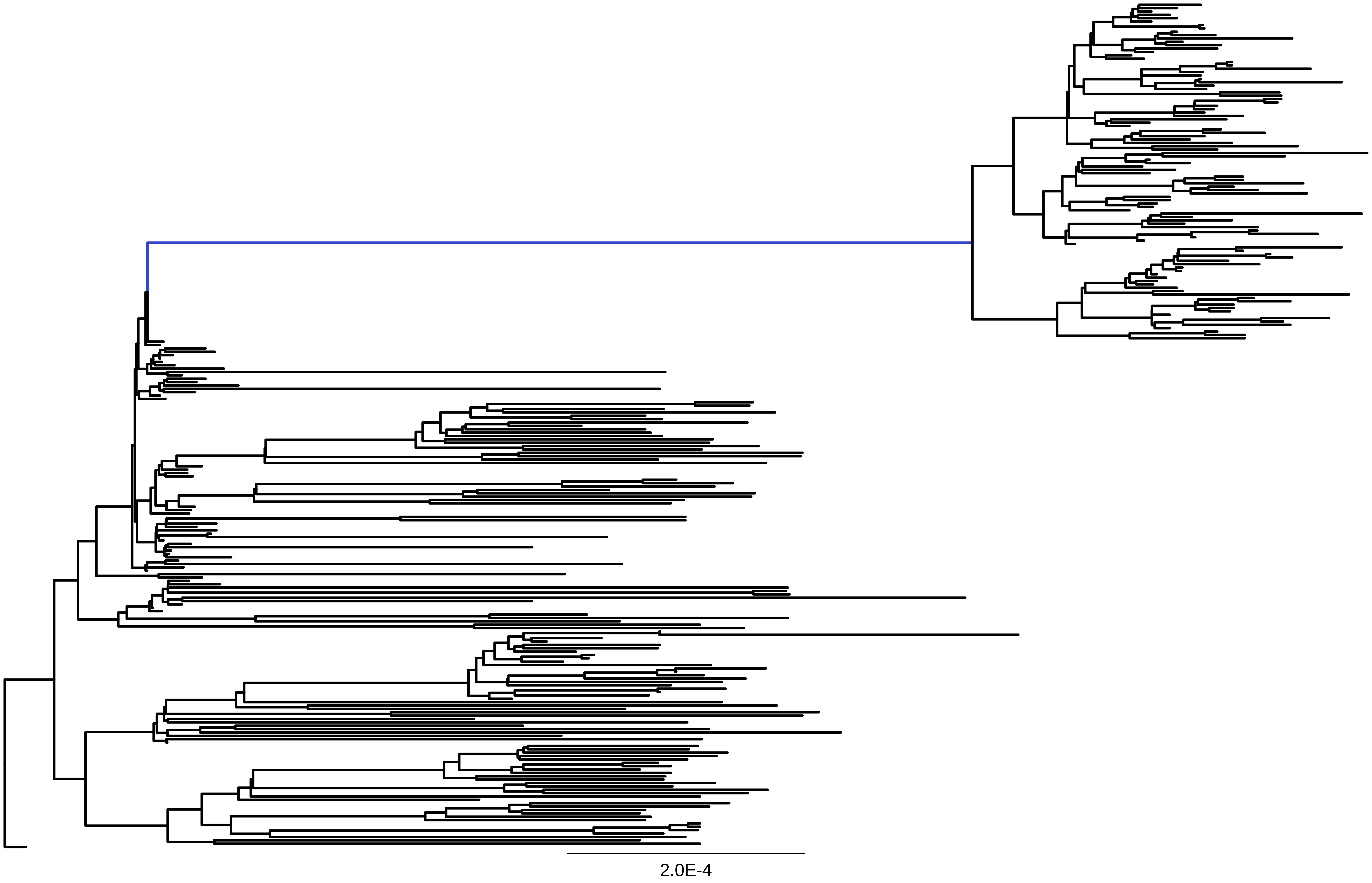

### Supp Fig 4

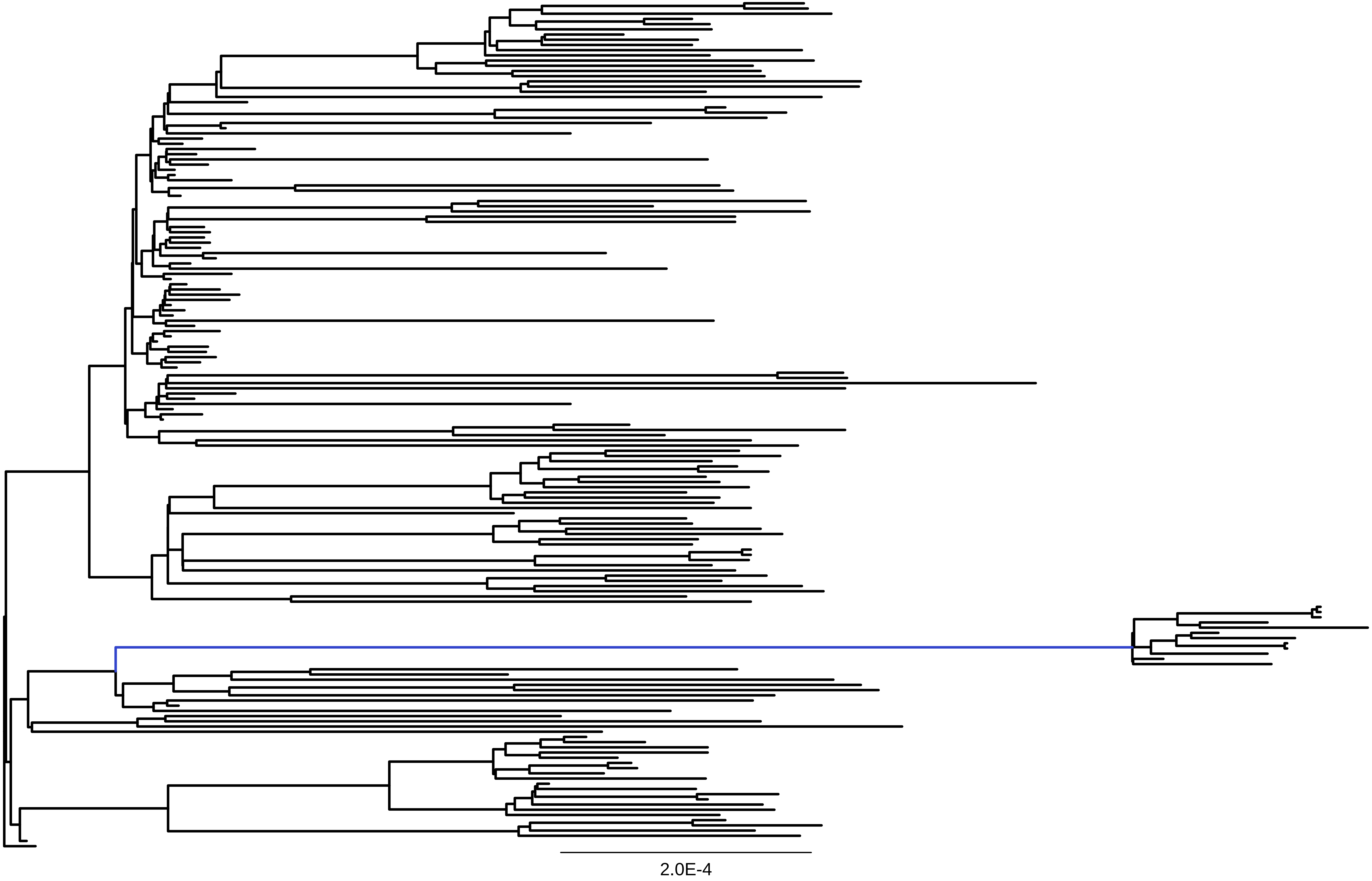

### Supp Fig 5

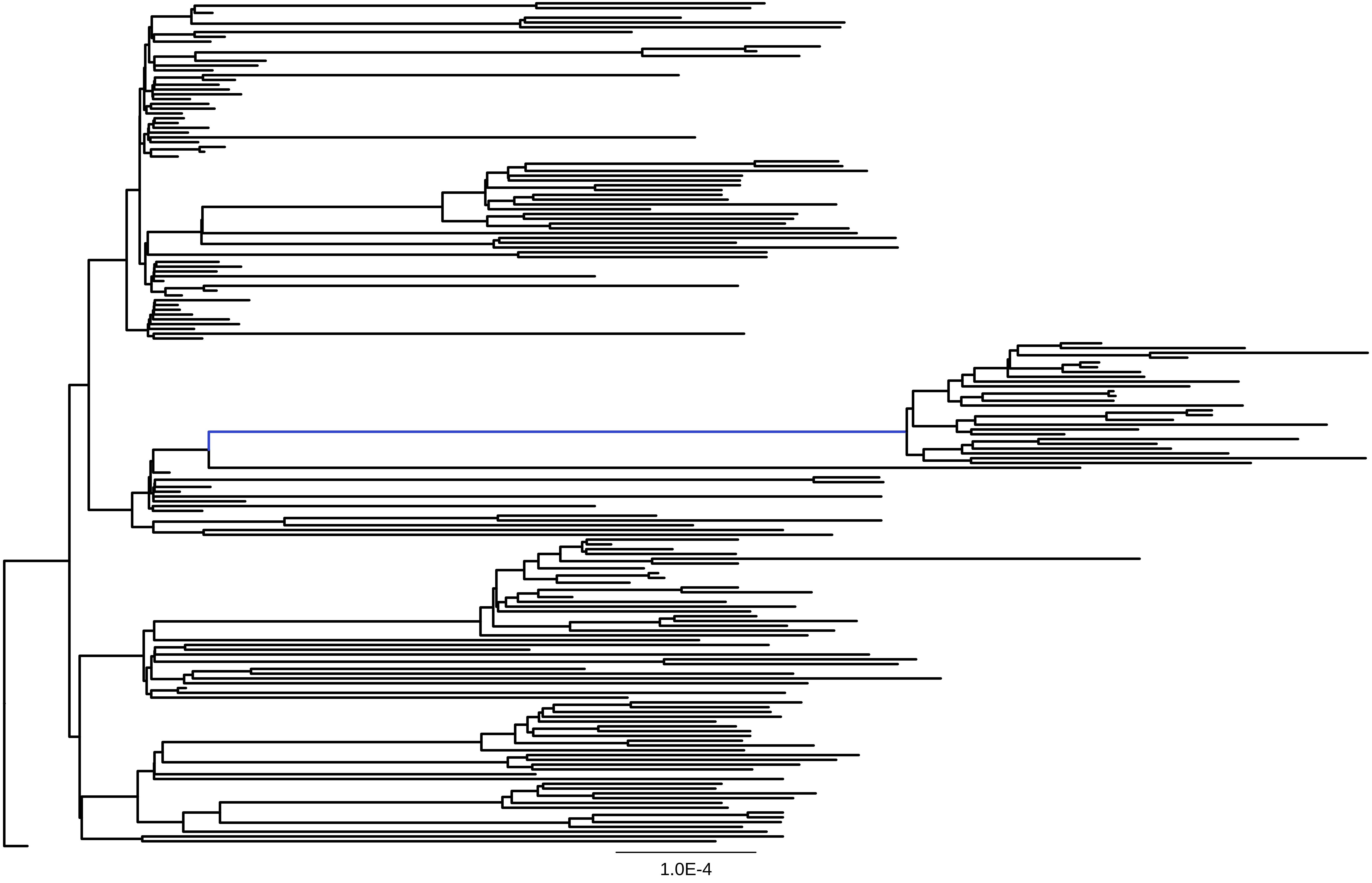

### Supp Fig 6

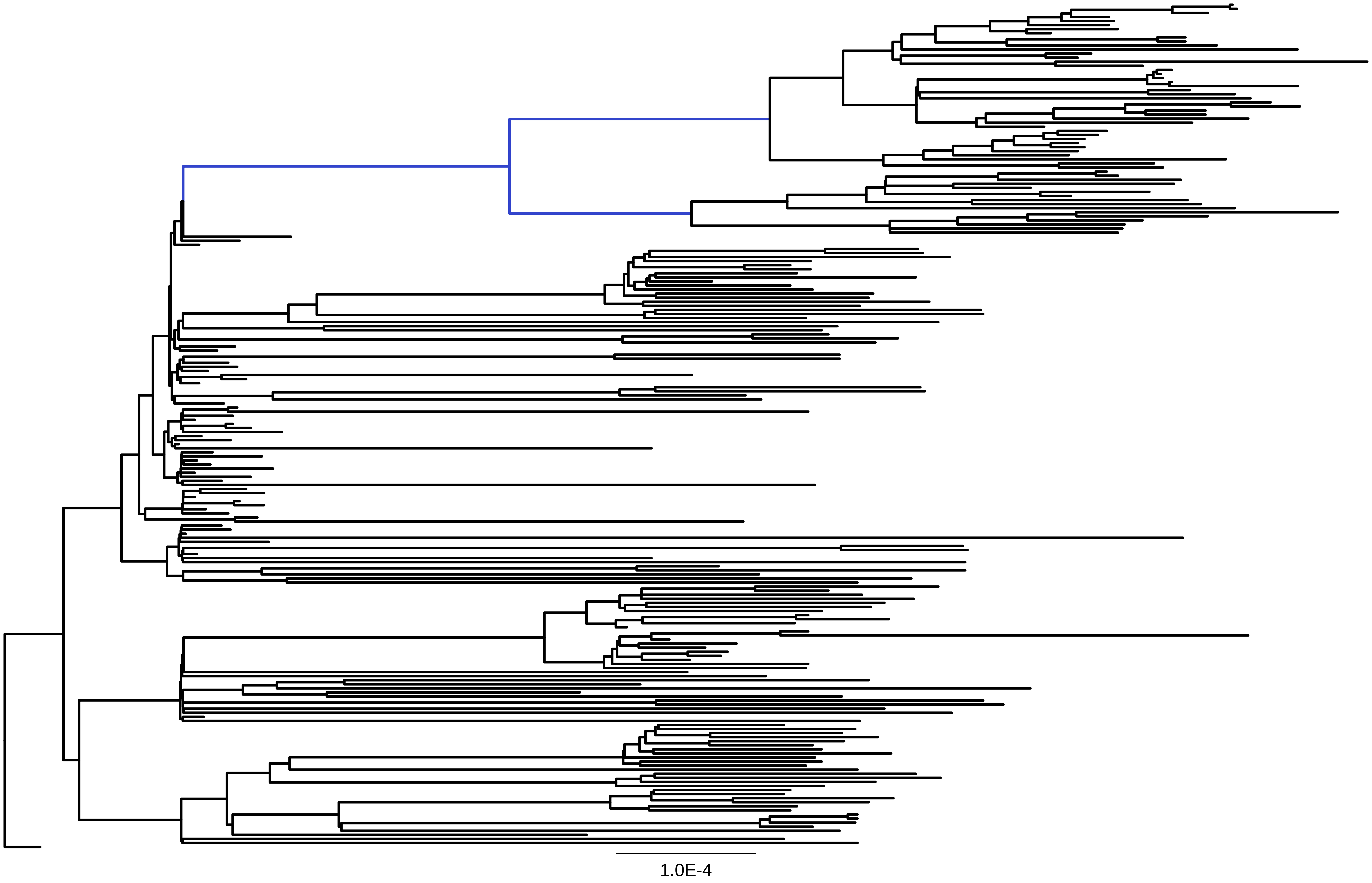

### Supp Fig 7

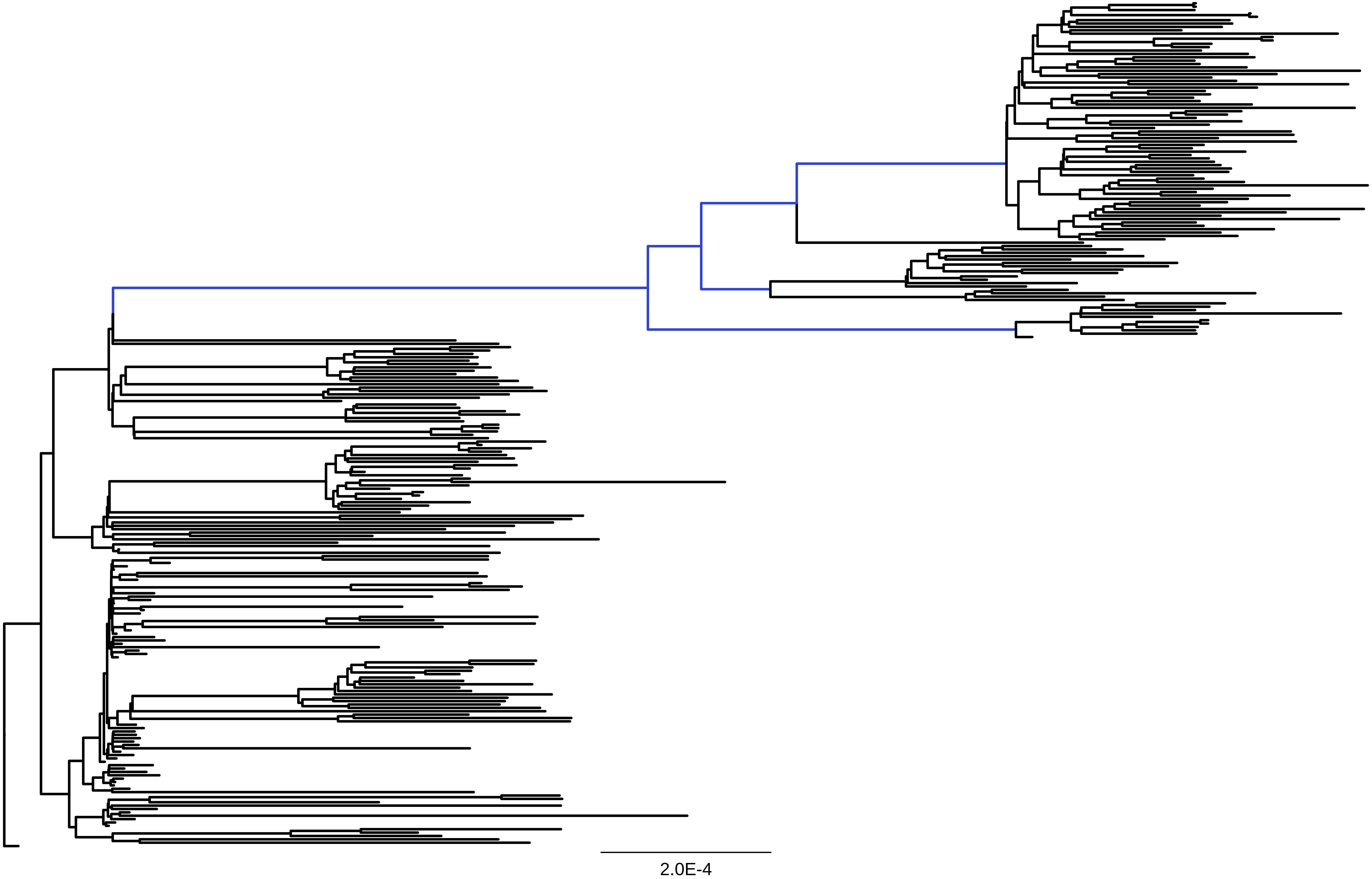

### Supp Fig 8

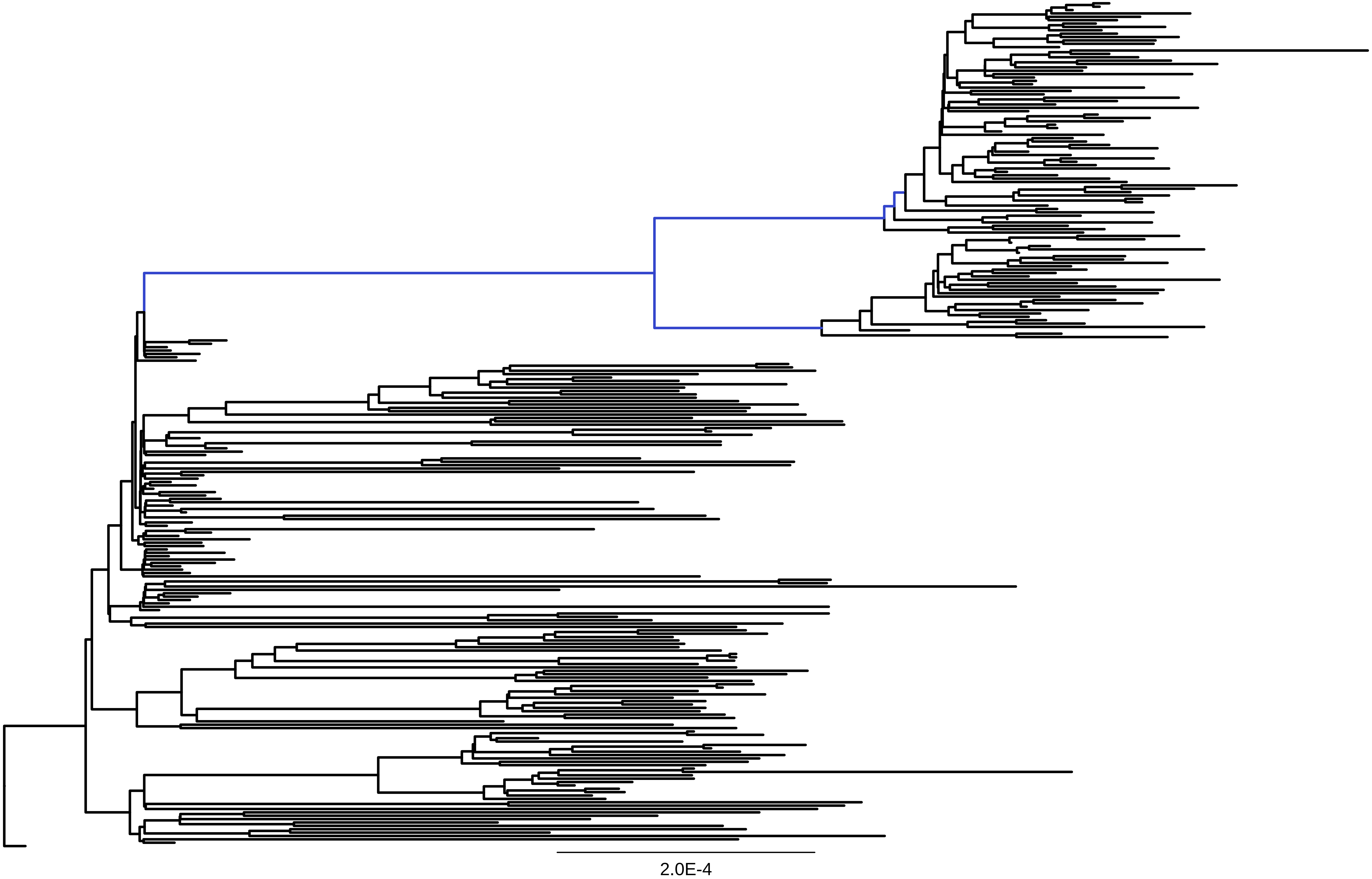

### Supp Fig 9

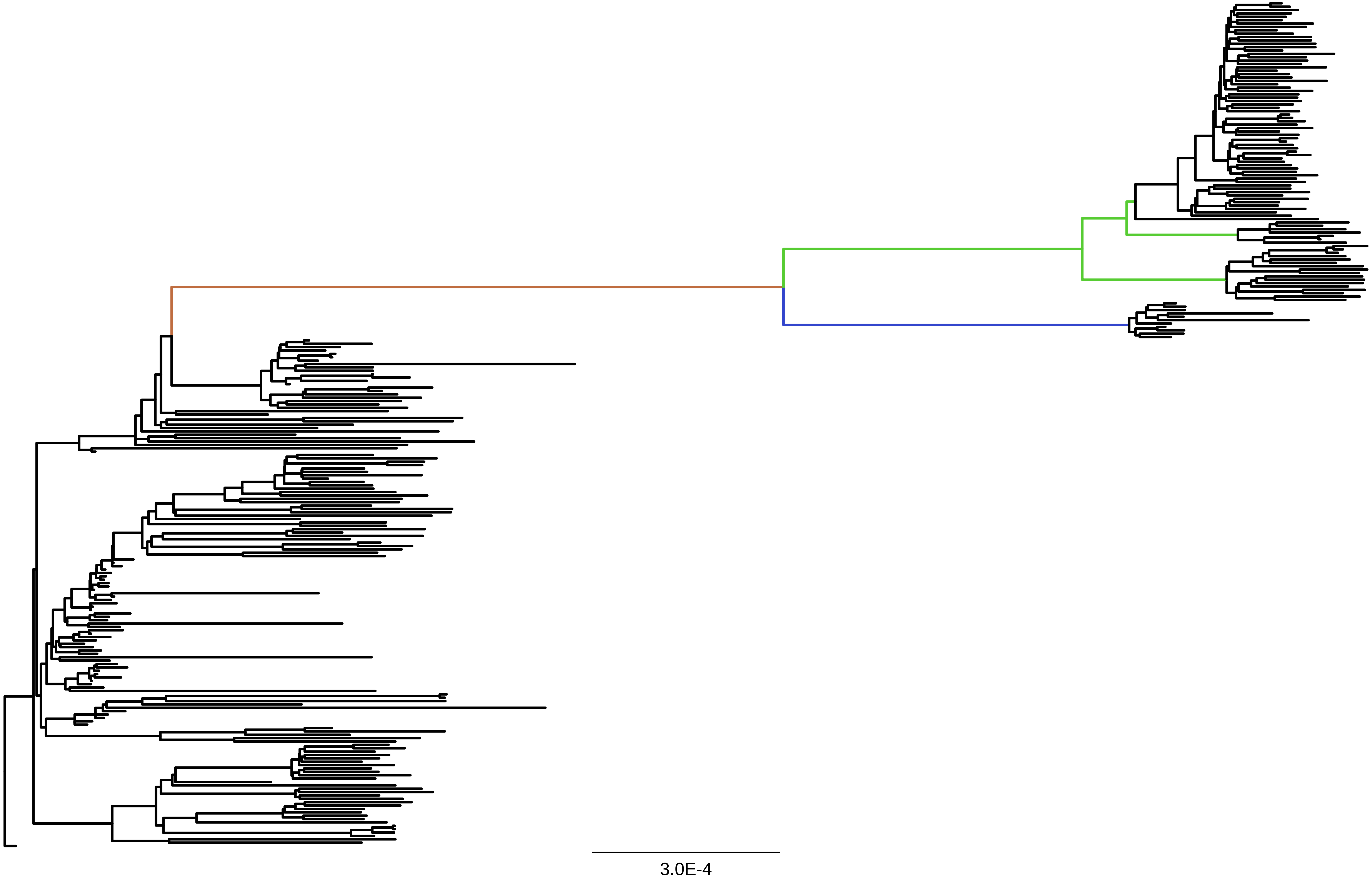

### Supp Fig 10

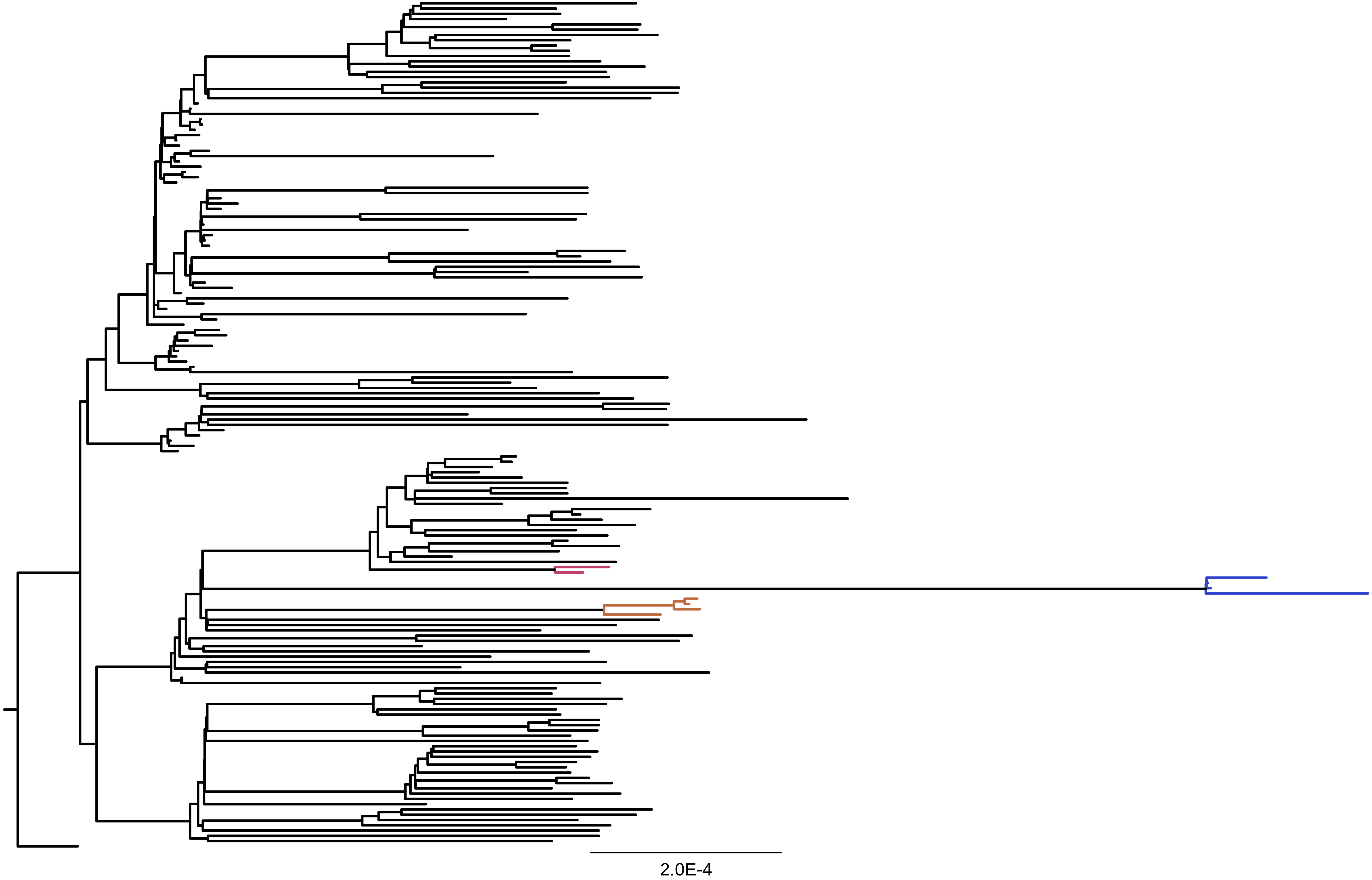

### Supp Fig 11

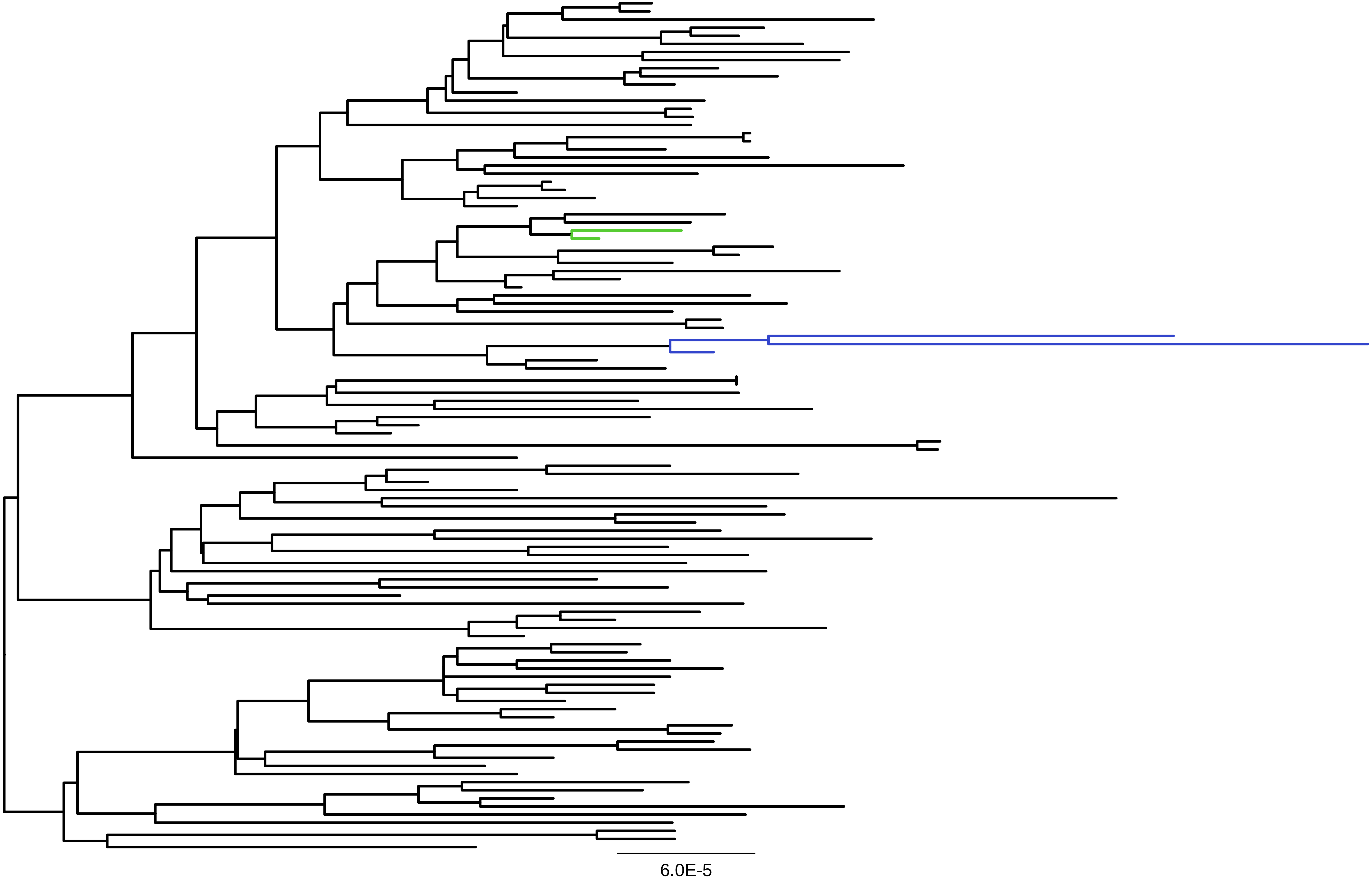

### Supp Fig 12

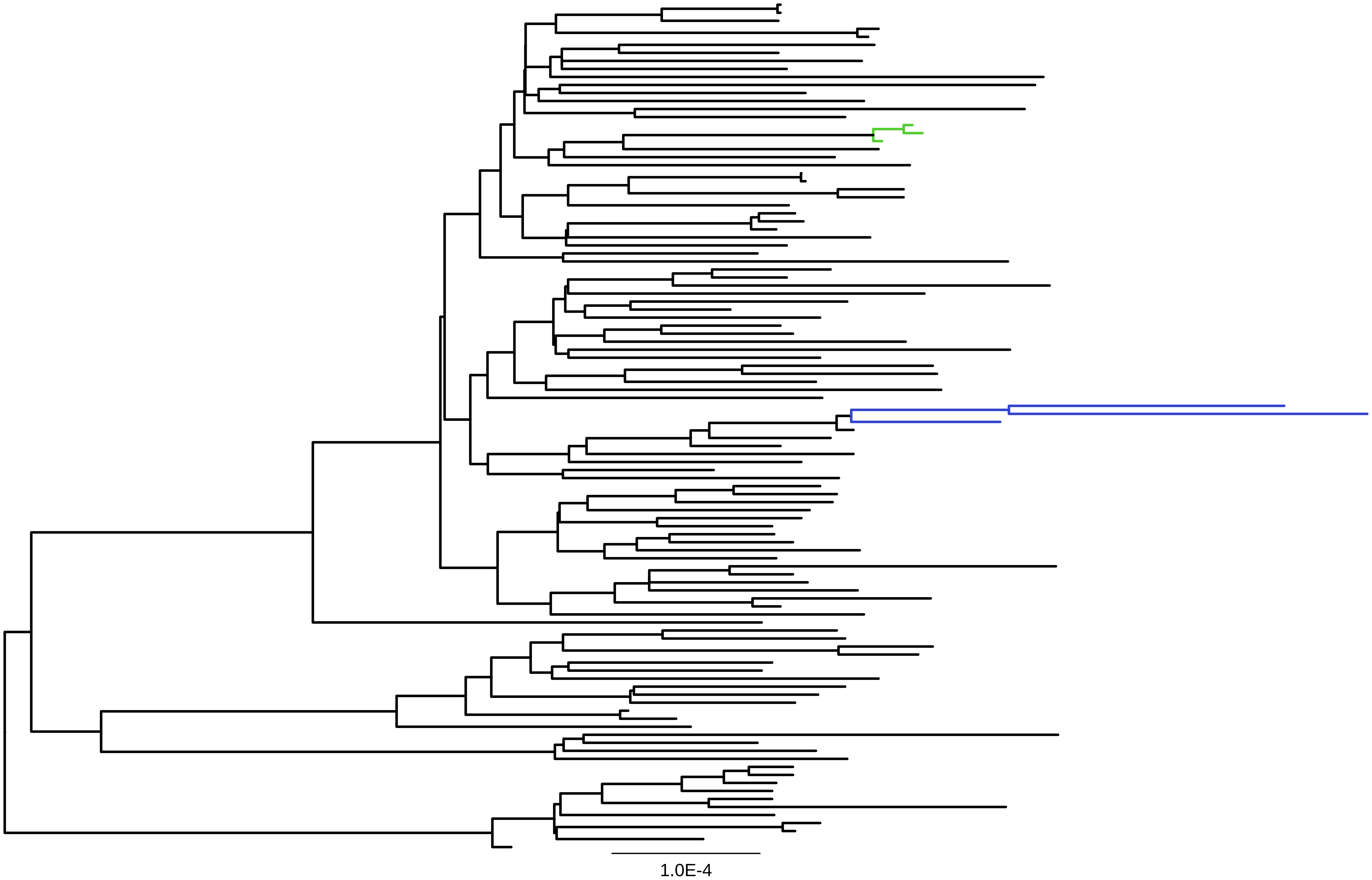

### Supp Fig 13

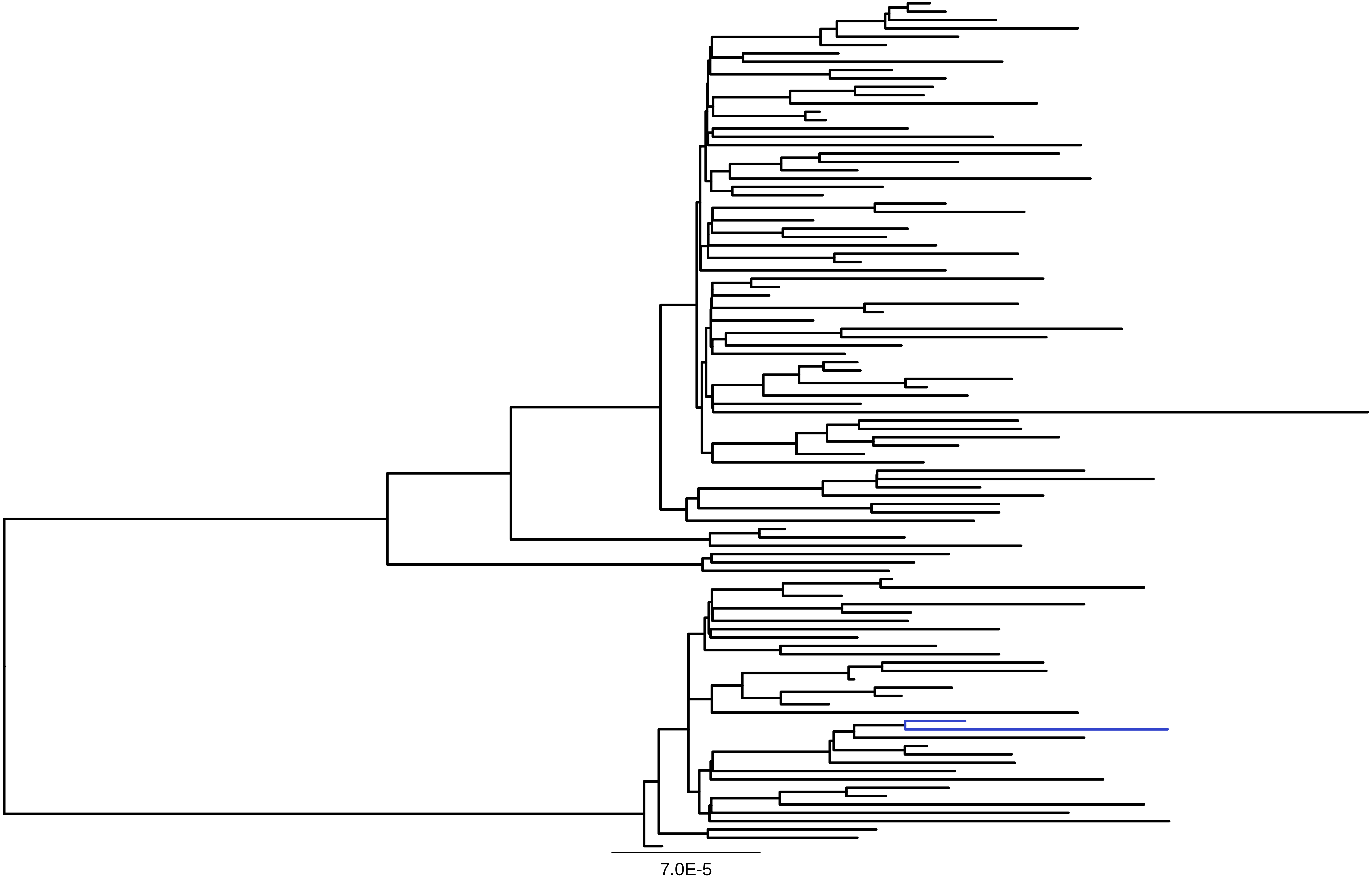

### Supp Fig 14

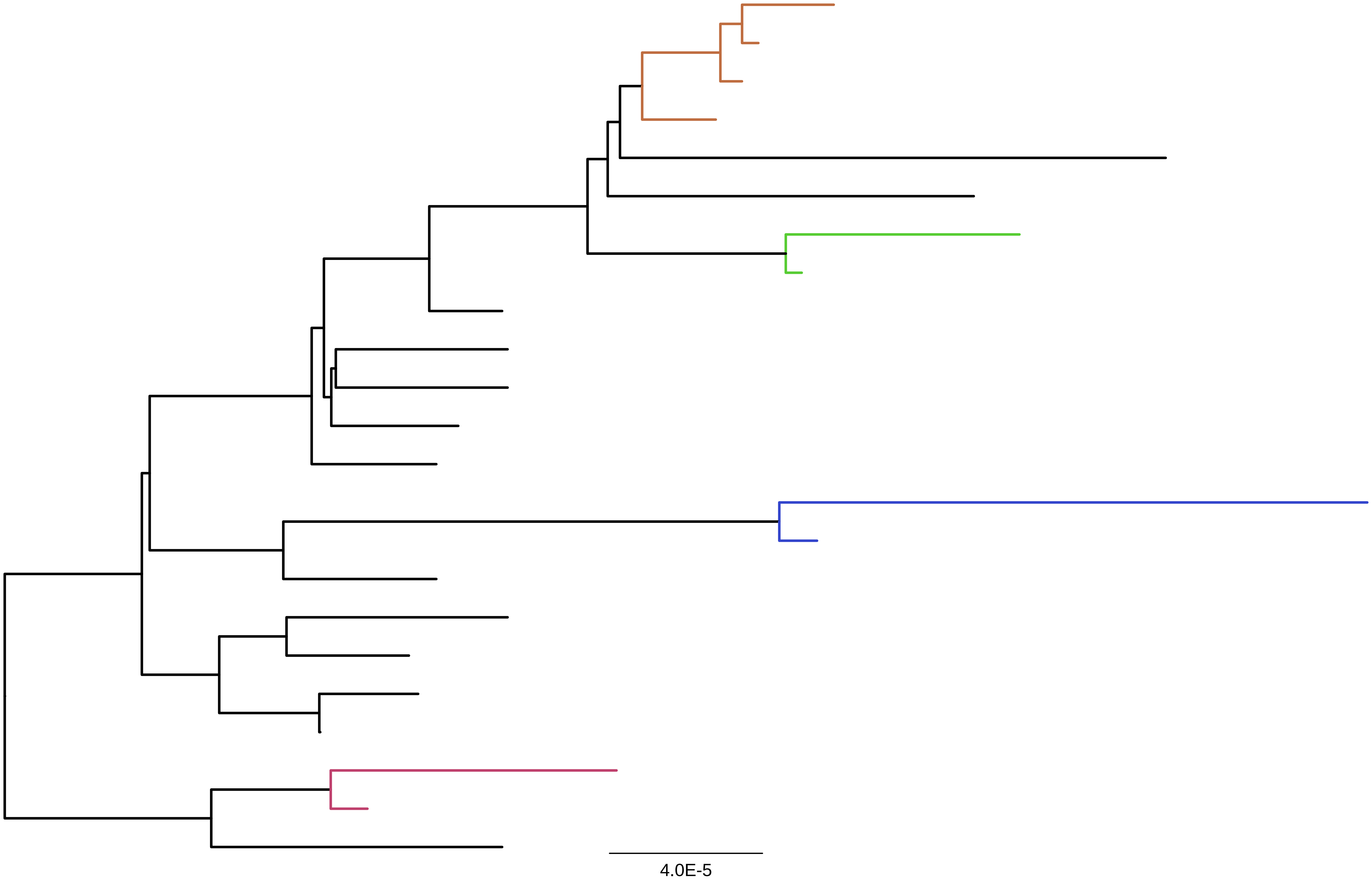

### Supp Fig 15

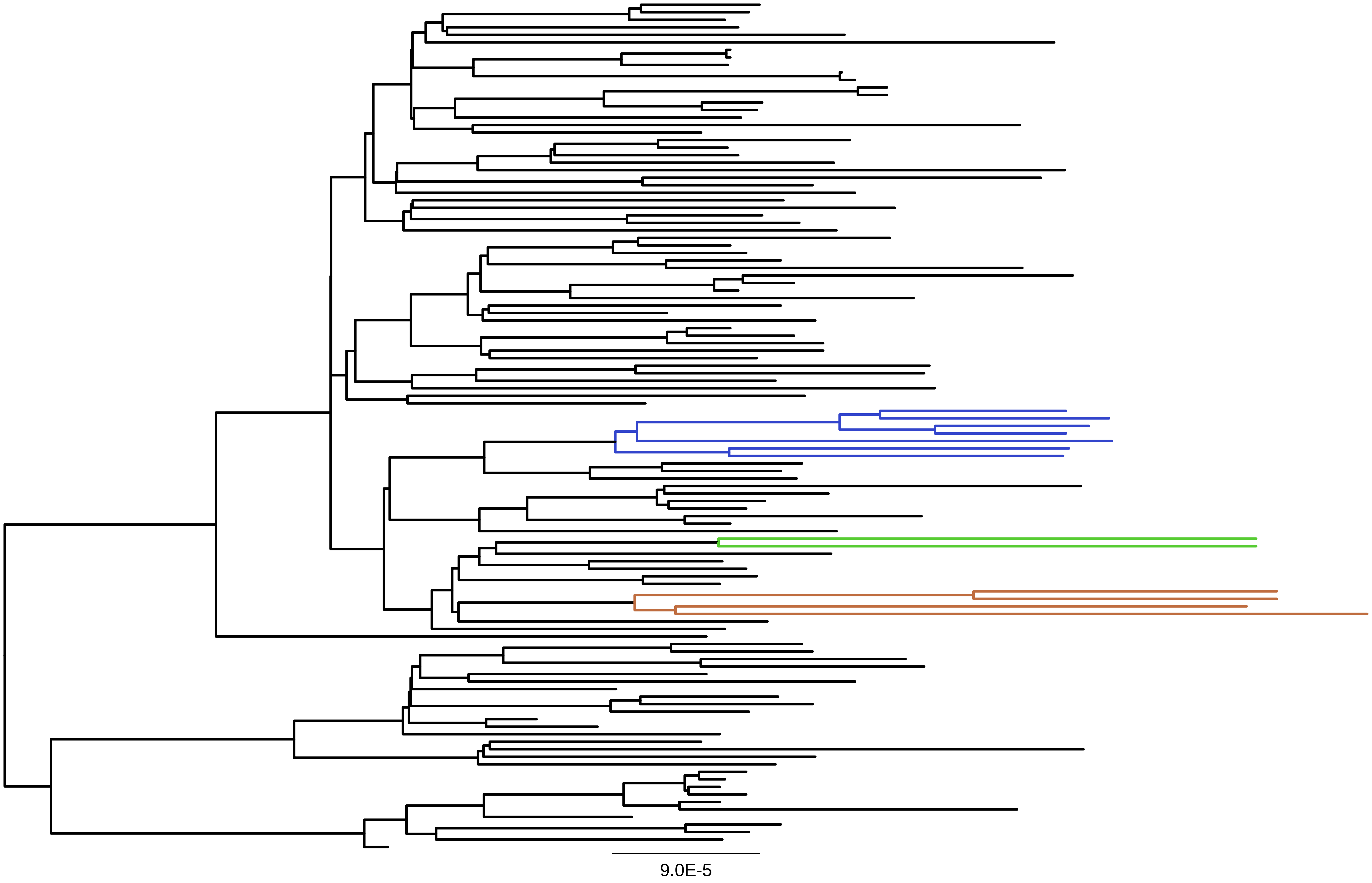

### Supp Fig 16

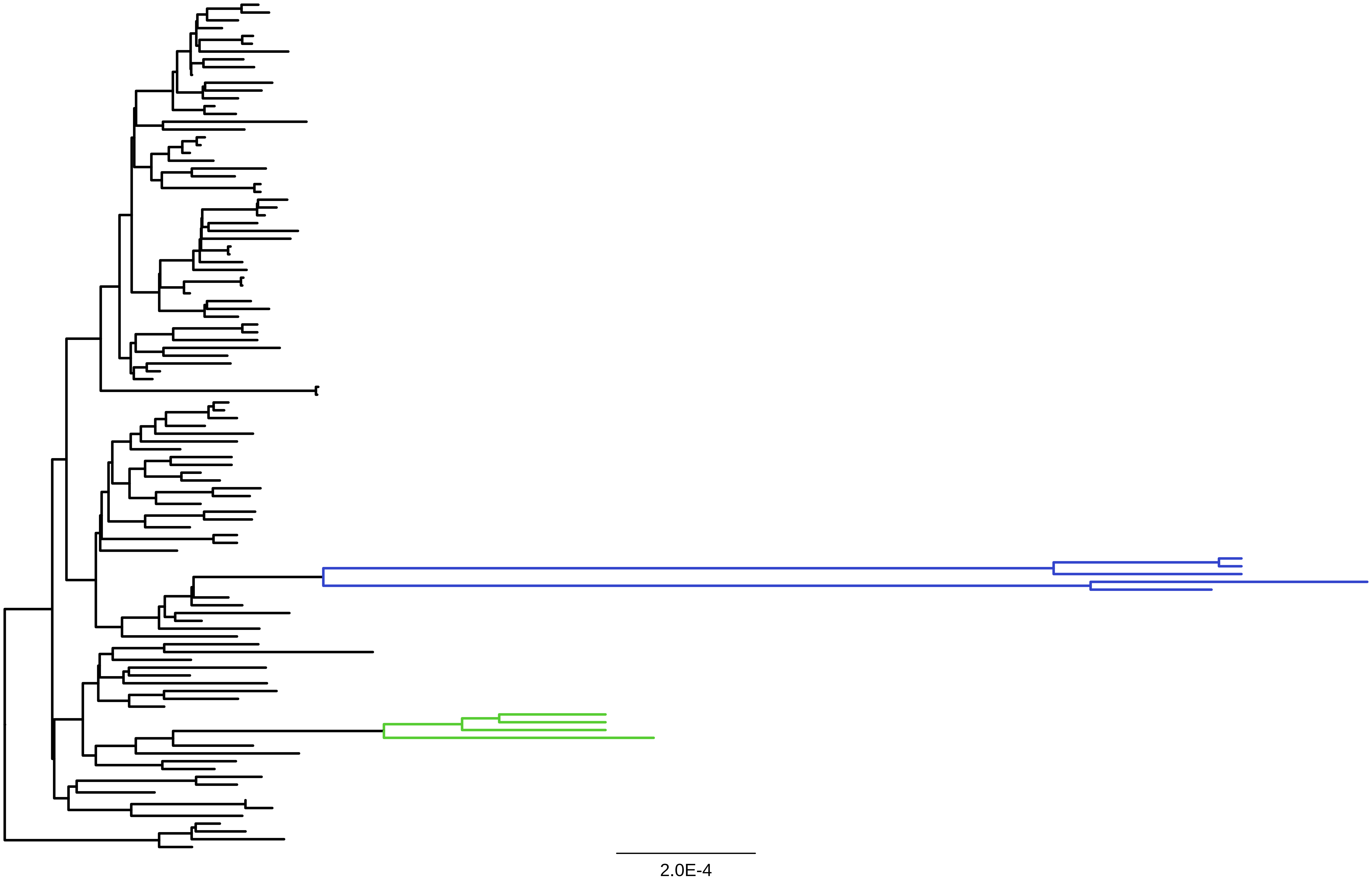
